# Understanding Tooth Agenesis: A Multi-omics Insight into MicroRNA Regulation

**DOI:** 10.1101/2024.09.02.24312916

**Authors:** Prashant Ranjan, Chandra Devi, Neha Verma, Rajesh Bansal, Vinay Kumar Srivastava, Parimal Das

## Abstract

This study reveals novel microRNAs (miRNAs) implicated in congenital tooth agenesis (CTA), a common dental anomaly with a complex genetic basis. Through a multi-omics approach combining bioinformatics, whole exome sequencing, metabolite analysis, and gene expression profiling, we identified eight key miRNAs potentially involved in tooth development. Among these, four miRNAs viz. miR-218-5p, miR-15b-5p, miR-200b-3p, and let-7a-3p were validated as significant regulators in CTA, marking their first investigation in blood samples from CTA patients. Our analysis revealed that these miRNAs play critical roles in odontogenesis, influencing essential signaling pathways, including *Wnt, FGF*, and *PI3* kinase pathways. Among these four, miR-218-5p and let-7a-3p emerged as key players in dental tissue morphogenesis, each contributing to previously unidentified networks crucial for tooth development.

This study highlights the potential of these miRNAs as non-invasive biomarkers for early CTA diagnosis and therapeutic targets. This is the first comprehensive investigation of these specific miRNAs in CTA, utilizing a multi-omics approach to offer fresh insights into miRNA-mediated mechanisms and their role in regulating dental anomalies. Our findings not only advance the understanding of the genetic regulation of tooth development but also pave the way for personalized approaches in managing dental anomalies. Further research is needed to validate these results and explore their clinical applications.

**Graphical Abstract:** 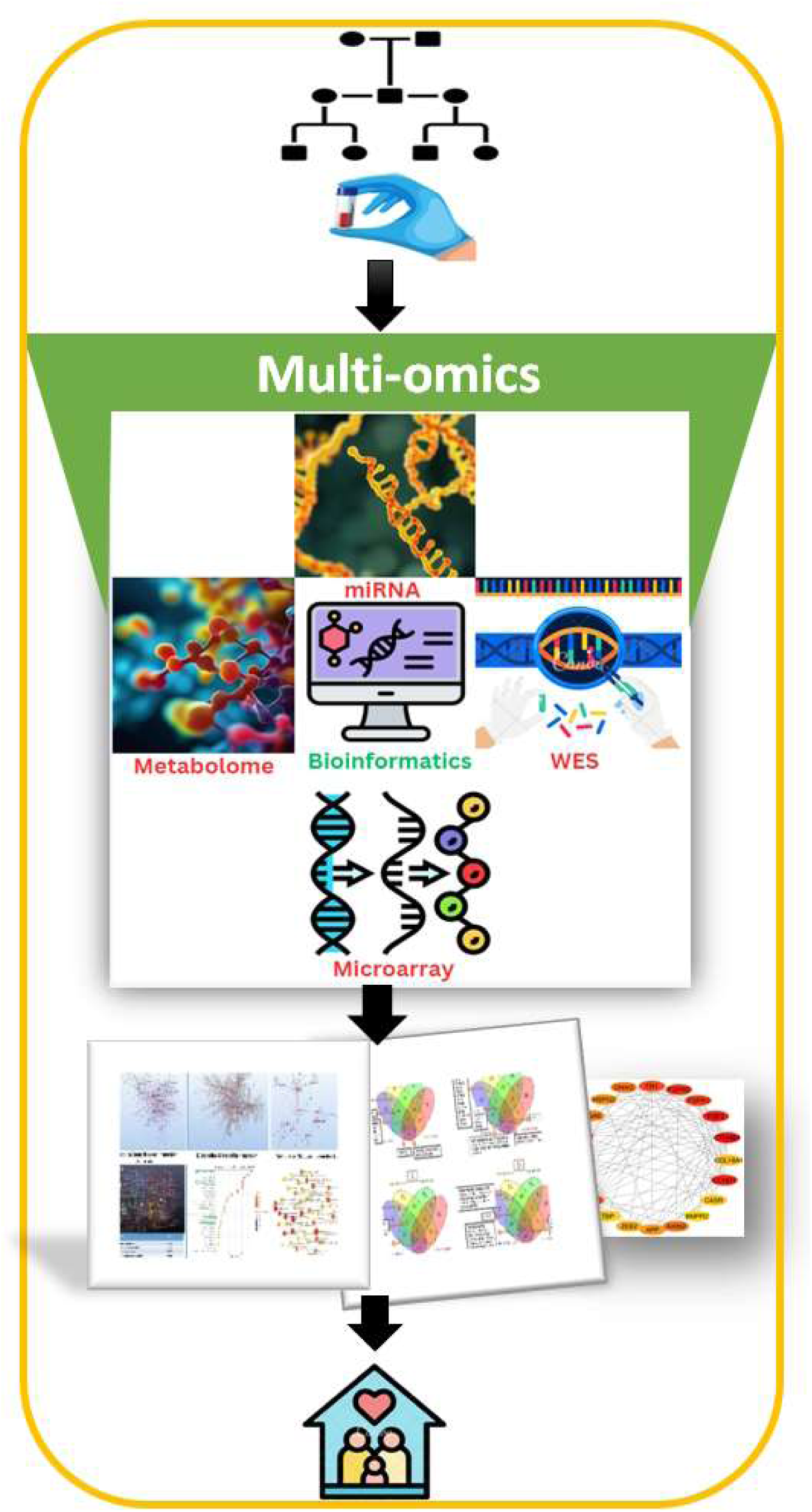

## 1. Introduction

Congenital Tooth Agenesis (CTA) is a developmental anomaly marked by the congenital absence of one or more teeth, significantly affecting oral health, function, and aesthetics. This condition ranges from hypodontia (missing up to six teeth) to oligodontia (missing six or more teeth), and anodontia (complete absence of teeth) (Azzaldeen et al., 2017). Effective management of CTA necessitates a thorough understanding of the intricate processes involved in tooth development, which hinge on interactions between the dental epithelium and mesenchyme (Pekka, 2009).

Tooth development begins with the thickening of the oral epithelium to form the dental lamina. Multiple dental placodes emerge within the dental lamina, initiating the formation of tooth germs. These placodes bud into the mesenchyme, inducing condensation and continuing to extend, shaping the tooth. The subsequent stages involve crown formation, which detaches the tooth from the oral epithelium, followed by root formation, which connects the teeth to nerves, blood vessels, and the alveolar bone, thereby initiating the eruption process (Matalová et al., 2015).

Numerous molecular pathways and genes are critical in this developmental process. Highly conserved pathways, such as *Wnt*/β-catenin, *BMP*, *FGF*, and Sonic Hedgehog (*SHH*), play pivotal roles and are tightly regulated throughout all stages of tooth development. In particular, the *Wnt*/β-catenin and *SHH* pathways are crucial for every transition step in odontogenesis (Hermans et al., 2021).

MicroRNAs (miRNAs) are small, non-coding RNAs that regulate gene expression post-transcriptionally by binding to the 3’ untranslated regions (3’ UTR) of target mRNAs, thereby inhibiting translation or promoting mRNA degradation. Recent research has highlighted the significant role of miRNAs in tooth development and agenesis (Giovannetti et al., 2024; Jin et al., 2017; Kural et al., 2024)

Disruptions in odontogenesis can result in various dental malformations, with dental agenesis being the most prevalent, affecting 0.15% to 16.2% of the population (Rakhshan, 2015). Additional anomalies include ectopic eruption and tooth impaction, both of which lead to malocclusion and have prevalence rates of 3.9% (Laganà et al., 2017) and 5.4% (De Ridder et al., 2022). Over the past decade, genetic variations or altered gene expression have linked molecular pathways such as *Wnt*, *TGF*, and *EDA*/*EDAR*/*NF-κB* to these conditions (Yu et al., 2019).

The role of miRNAs in dental anomalies remains incompletely understood. To date, studies on miRNAs in CTA have primarily focused on dental tissues such as dental pulp, periodontal ligament, and developing tooth germs (Sera & Zur Nieden, 2017). However, research on miRNA analysis in blood samples in the context of CTA is limited. The potential advantage of using blood samples lies in their minimally invasive collection and their ability to provide a systemic view of miRNA expression profiles. Blood-based miRNA biomarkers could enable early diagnosis, monitoring, and personalized treatment strategies for dental anomalies, enhancing the overall management of conditions like CTA. Future research should focus on elucidating the complete miRNA-mRNA networks, exploring their potential as diagnostic and therapeutic tools, and advancing regenerative strategies for dental anomalies. Understanding miRNA-mediated pathways holds promise for innovative treatments and improved management of CTA.

In this study, we identified potential miRNAs involved in tooth development through comprehensive literature mining and bioinformatics analysis. We compiled a list of miRNAs implicated in CTA and other dental-related conditions, particularly those affecting epithelial and mesenchymal cell functions. Based on our bioinformatics analysis, we selected eight key miRNAs for further investigation, marking the first time these miRNAs have been analyzed in blood samples from patients with CTA. We developed multi-omics gene panels and cross-validated the association of four significant miRNAs, identified through RT-qPCR analysis in blood samples from CTA patients, with whole exome sequencing (WES), metabolite, and gene expression panels. This study is the first to investigate these miRNAs in the context of CTA. Following this, we conducted pathway and biological function analysis to elucidate the role of these miRNAs in regulating the ontogenesis process, particularly concerning dental anomalies such as agenesis and impaction.

## 2. MATERIALS AND METHODS

### 2.1 Study Population

This study involved participants diagnosed with CTA. Inclusion criteria were: (1) confirmed diagnosis of congenital CTA, (2) age between 18 and 35 years, (3) availability of comprehensive dental records and radiographs, and (4) consent to participate, provided by the participants or their legal guardians. Exclusion criteria were: (1) tooth loss due to reasons other than congenital agenesis, (2) incomplete dental documentation, or (3) prior orthodontic treatment. A total of 10 patients and 5 control samples were obtained from the Department of Dentistry, Oral Surgery, and Medicine at the Institute of Medical Sciences, India. The patients were scheduled for clinical examinations and dental implant procedures. Five well-matched controls, aged 25 to 35, were selected to closely resemble the patients.

### 2.2 Data Collection

Each participant’s dental records and radiographs were carefully reviewed to confirm the diagnosis and assess the extent of CTA. Clinical examinations were conducted by qualified dental professionals to ensure accuracy and consistency in data collection.

### 2.3 Blood Collection, Storage, and RNA Isolation

Peripheral venous blood samples (1.5 mL) were collected using a 5 mL heparinized syringe. The samples were transported to the laboratory within 60 minutes of collection, maintained in a cooling box to preserve stability. RNA was extracted from 200 µL of blood using the Trizol reagent (Sigma-Aldrich, Catalog No. T9424). The concentration and quality of the extracted RNA were assessed using a NanoDrop spectrophotometer (Thermo Scientific, USA).

### 2.4 MicroRNA Panel Selection

#### 2.4.1 Literature Review

A comprehensive literature search was conducted on PubMed using the keywords “miRNA and tooth development,” yielding 121 research articles published between 2008 and 2023 (Supplimentary File1).

#### 2.4.2 Identification of miRNAs

The identified articles were reviewed to compile a list of miRNAs associated with tooth development. After eliminating duplicates, a refined list of relevant miRNAs was created (Supplimentary File1).

#### 2.4.3 Retrieval of Target Genes

The miRDB database was accessed to investigate the regulatory network of the selected miRNAs. A target score threshold of 90 or higher was applied to select miRNA target genes, resulting in a list of miRNAs crucial to CTA (Supplimentary File1)..

#### 2.4.4 Collection of Genes Associated with CTA

A comprehensive list of 133 genes associated with congenital tooth agenesis (CTA) was compiled by searching the GeneCard and OMIM databases (Supplimentary File2_Gene panel).

#### 2.4.5 Mapping miRNA-Target Gene Interactions

Interactions between miRNAs and their target genes within the CTA context were identified by cross-referencing miRNA target genes with CTA-associated genes. This analysis led to the identification of 58 miRNAs and their corresponding target genes for further investigation (Supplimentary File1).

#### 2.4.6 Network Analysis

The interactions between miRNAs and their target genes were visualized and analyzed using Cytoscape version 3.10.1 (Doncheva et al., 2018). This analysis identified 25 miRNAs targeting a single gene, 13 miRNAs targeting two genes, eight miRNAs associated with three genes, and 13 miRNAs interacting with more than three genes. The disease associations of these miRNAs were determined using the miR2Disease database **(Figure 1)** (Jiang et al., 2009).

**Figure 1.**
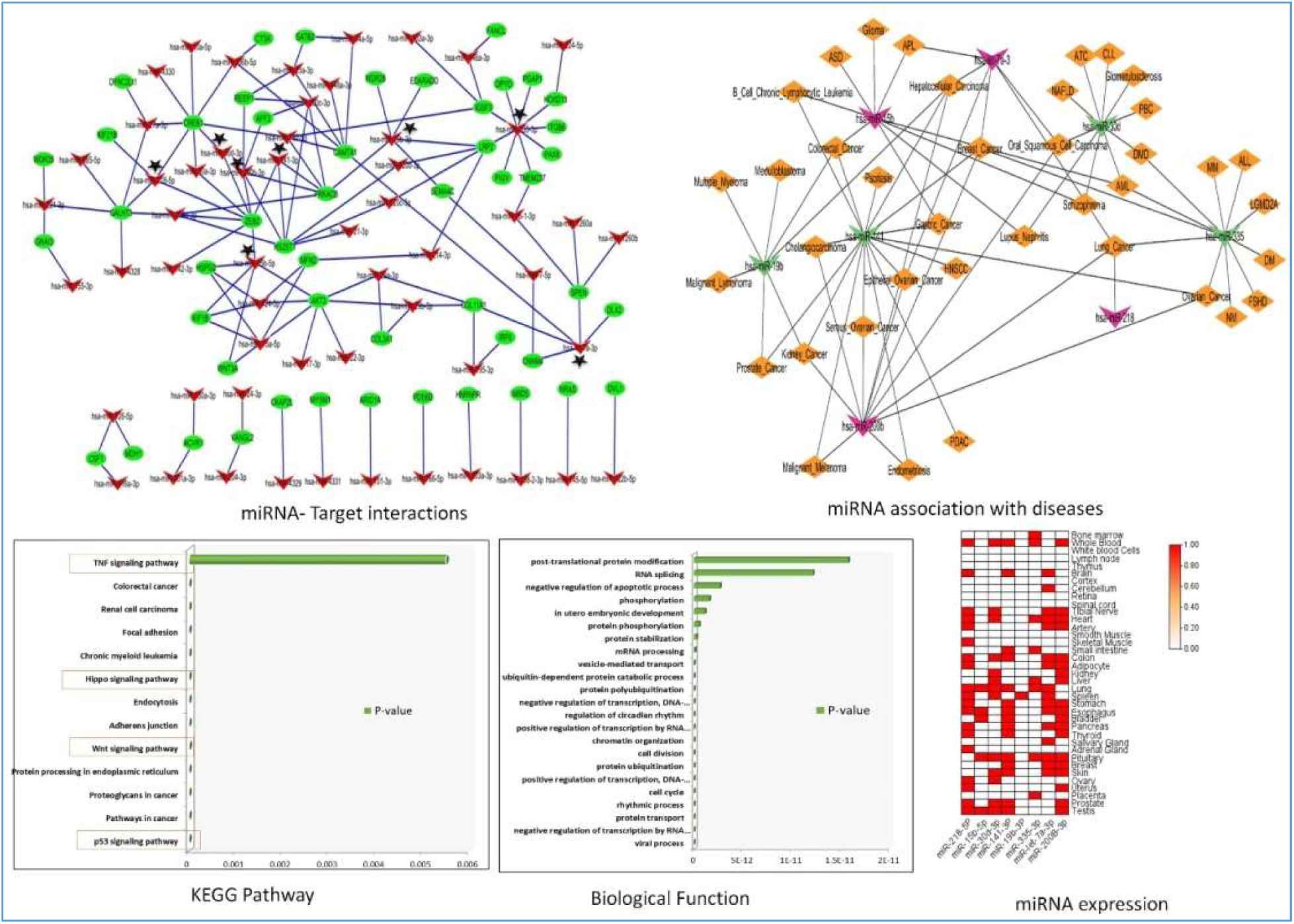
Selection and Functional Analysis of 8 miRNAs for *In Vitro* Studies Related to Tooth Development. This figure illustrates the selection process of 8 miRNAs (miR-335-3p, miR-15b, miR-30d, let-7a, miR-218, miR-141, miR-19b, and miR-200b) for in vitro studies. Selection was based on network analysis, emphasizing miRNAs with extensive target gene connections and those targeting similar genes. The network analysis highlights their significant roles in regulatory networks. Although these miRNAs are associated with various diseases according to the miR2Disease database, none have established links to dental diseases. KEGG and biological function analyses support their involvement in tooth development, and a heat map shows their expression across different human tissues.

#### 2.4.7 Overlapping Gene Network Construction

To identify genes targeted by multiple miRNAs, a network of overlapping genes obtained after multi-omics analysis was constructed. Nodes represented individual genes, while edges depicted miRNA-gene interactions, emphasizing shared targets across different miRNAs. These shared genes were further analyzed using the STRING database (Mering et al., 2003) to assess protein-protein interactions (PPI), which were visualized in Cytoscape version 3.10.1(Doncheva et al., 2018). The CytoHubba plugin (Chin et al., 2014) was used to identify the top 20 hub genes based on the Maximum Clique Centrality (MCC) scoring method, highlighting their importance within the network.

#### 2.4.8 Pathway Analysis for miRNAs

Pathway analysis was performed using the miRPath V4.0 tool (Diana Tool)(Tastsoglou et al., 2023) to gain insights into the biological significance of the 13 miRNAs that showed interactions with three or more genes. Various pathway resources, such as KEGG and GO Biological Process keywords, were used to understand the functional roles of these miRNAs.

### 2.5 miRNA Stem-Loop RT-qPCR Analysis

Eight miRNAs of particular relevance were selected for further functional studies. Stem-loop RT-qPCR primers for these miRNAs were designed using the Srna Primer DB online tool (Xie et al., 2019) (Supplimentary File1). The nucleotide sequences of these miRNAs were retrieved from the miRBase database. Additionally, U6 small nuclear RNA was included as a potential endogenous control in the panel.

### 2.6 Reverse Transcription and Quantitative RT-PCR

Reverse transcription of miRNA samples was performed using the Revert Aid First Strand cDNA Synthesis Kit (Thermo Scientific, Catalog #K1622, USA). MiRNA-specific stem-loop reverse transcription primers, as listed in Supplementary Table X, were added to the reaction mixture. Quantitative PCR (qRT-PCR) was conducted using the Applied Biosystems QuantStudio 6 Flex Real-Time PCR System (Thermo Scientific, USA). The reaction mixture (12.5 μL) included 1 μL of pre-amplified cDNA (diluted 1:5), 6.25 μL of Maxima SYBR Green/ROX qPCR Master Mix (2X, Thermo Scientific, Catalog #K0221, USA), 1 μL of 10 μM miRNA-specific forward primer, 1 μL of 10 μM reverse primer, and nuclease-free water.

### 2.7 WES Panel

WES was conducted utilizing the Illumina Next-Generation Sequencing platform, focusing on approximately 30Mb of the human exome. This approach encompassed nearly 99% of the coding regions as defined by CCDS and RefSeq. Sequencing achieved a mean depth of 80-100X, with over 90% of the targeted regions covered at a minimum depth of 20X. To ensure data quality, duplicate reads were eliminated, and base quality recalibration was performed. The reads were mapped to the human reference genome GRCh38. Variant calling followed the GATK best practices, and subsequent variant annotation was carried out using databases such as OMIM, GWAS, gnomAD, and 1000 Genomes. Filtration of the annotated variants was based on several criteria, including a gnomAD allele frequency ≤ 0.01 or unavailable, a minimum read depth of 20, pathogenicity, and predicted functional consequences. Upon confirming the presence of the mutant variant in the candidate gene with a minor allele frequency (MAF) of ≤ 0.01, the MAF threshold was subsequently adjusted to ≤ 0.20 (Supplimentary File2_Gene panel).

This broader range was applied to include all associated genes for further multi-omics analysis. We employed a stepwise filtering strategy to identify mutations in our candidate gene from an initial pool of 61,767 genetic variants. First, we filtered variants based on a minor allele frequency (MAF) threshold of ≤0.01, resulting in 2,900 rare variants. We then restricted the analysis to variants within 20 base pairs of exon boundaries, including exonic, splice-site, and non-synonymous variants, reducing the set to 1,334. Variants with a total read depth exceeding 20 were prioritized, leaving 1,109 variants, of which 1,058 met standard quality criteria. To further refine the selection, we applied a threshold CADD score of ≥20 (or where unavailable), narrowing the pool to 391 variants. By applying ACMG guidelines and excluding benign variants, 330 potentially pathogenic variants were identified. Finally, after assessing for high predicted severity and homozygosity, 79 variants remained, including 2 homozygous variants.

### 2.8 Microarray Gene Expression Panel

Gene expression data (GES56486) was obtained from the GEO database(Barrett et al., 2005) for patients with tooth agenesis, characterized by clinical manifestations such as nail dystrophy or loss, marginal palmoplantar keratoderma, hypodontia (missing ≤5 teeth), enamel hypoplasia, oral hyperpigmentation, and dysphagia. The raw data were normalized using the RMA method in R. Differentially expressed genes were identified using the limma package in R (version 3.5.2), with a log2 fold change (Log2FC) threshold greater than 1 for upregulated genes and less than −1 for downregulated genes. A p-value threshold of < 0.05 was applied for statistical significance (Supplimentary File2_Gene panel).

### 2.9 Metabolite Gene-Regulated Panel

For our study, we compiled a list of metabolites involved in tooth development and related processes such as bone development using literature sources (Yu et al., 2021) and the Human Metabolome Database (HMDB). Keywords like “Tooth,” “Tooth Enamel,” “Bone,” “Ectodermal Dysplasia,” “Cleidocranial Dysplasia,” and “Osteoblast” were used to retrieve relevant metabolites for further analysis. Metabolomic data was processed and analyzed using MetaboAnalyst 6.0, which included pathway enrichment and impact analysis based on databases like KEGG and SMPDB. Significant pathways were identified based on p-values, impact scores, and enrichment scores.

Key metabolic pathways associated with tooth development, such as Vitamin D3/Calcitriol, Vitamin A, collagen synthesis (L-Proline, 4-Hydroxyproline), Citric Acid Cycle, prostaglandin pathways, testosterone, and thyroid hormones, were identified and included in further analysis. These pathways were linked to processes like enamel and dentin mineralization, inflammation, tissue remodeling, and tooth eruption (Botelho et al., 2020; Costello et al., 2012; Ern et al., 2019; Goldberg et al., 2011; Morkmued et al., 2017; ROBERTS, 2020; Vucic et al., 2017). Further analysis highlighted pathways potentially involved in tooth agenesis, including arginine and proline metabolism, glycine and serine metabolism, purine metabolism, and thyroid hormone pathways, which were integrated into a gene panel for further investigation (Supplimentary File2_Gene panel)(Andersson, 2018; Andujar et al., 1991; Karna et al., 2020; Shapiro, 2001; Vucic et al., 2017).

Finally, we used keywords from these pathways to search the OMIM database, identifying genes associated with the metabolic pathways of interest. This gene panel will facilitate further exploration of the molecular mechanisms underlying tooth development and related conditions.

### 2.10 Miscellaneous

The PANTHER (Protein Analysis Through Evolutionary Relationships) tool (Mi et al., 2017) was used to identify pathways and biological functions of genes obtained through combined analysis of multiple omics data, including WES, microarray, metabolome analysis, and miRNA target panel. Venn diagrams generated using Venny 2.0 (Oliveros, 2007) were used to compare gene panels, highlighting shared and unique genes across different omics platforms. This comparison helped identify key genes consistently implicated across datasets.

### 2.11 Statistical Analysis

Statistical analyses were performed using GraphPad Prism (Version 8.0.2; GraphPad Software, San Diego, USA) (Swift, 1997). Data were expressed as mean ± SD. Comparisons between groups were performed using Student’s t-test. A p-value of less than 0.05 was considered statistically significant.

## 3. Results

### 3.1 Differential Expression of miRNA

We analyzed the differential expression of miRNAs in a total of 15 samples, including 5 control samples and 10 samples from patients with congenital tooth agenesis (CTA). The initial selection of 8 miRNAs (miR-335-3p, miR-15b, miR-30d, let-7a, miR-218, miR-141, miR-19b, and miR-200b) was based on an extensive literature review followed by in silico analysis. These miRNAs were chosen for in vitro studies due to their extensive connections with target genes and the fact that multiple miRNAs target similar genes within regulatory networks.

Network analysis revealed the significant roles of these miRNAs in various diseases, as identified by the miR2Disease database; however, none had previously been linked to dental diseases. KEGG pathway and biological function analyses further supported their involvement in tooth development, with a heat map illustrating their expression across different human tissues.

All samples exhibited detectable levels of expression for the selected miRNAs. The boxplot in (Figure 2) demonstrates the differential expression of these miRNAs, represented by dCt values and corresponding p-values. Our analysis identified significant upregulation of miR-218-5p, miR-15b-5p, and miR-200b-3p, with p-values of 0.0123, 0.0001, and 0.0001, respectively. Conversely, miR-let-7a-3p was significantly downregulated (p-value = 0.0054). miR-19b-3p showed no major change in expression (p-value = 0.1786). miR-30d-3p was upregulated but not significantly (p-value = 0.09). miR-141-3p and miR-335-3p did not exhibit significant changes in expression, with p-values of 0.7297 and 0.805, respectively (Figure 2).

**Figure 2.**
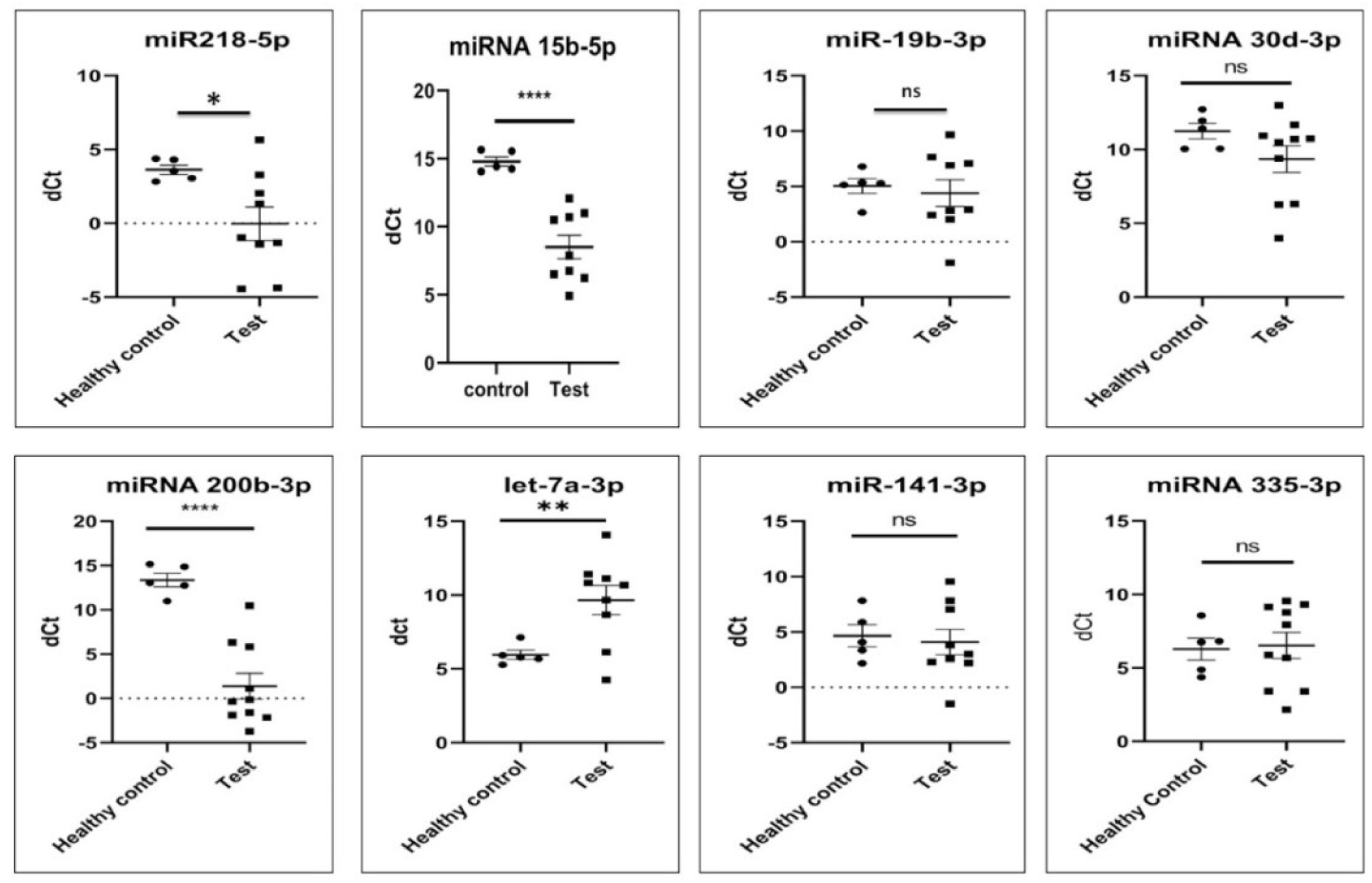
Whisker Plot Showing the ΔCt Value Expression in Different miRNAs. This figure presents a Whisker plot of the ΔCt values for the expression levels of different miRNAs. The ΔCt value, representing the difference in cycle threshold (Ct) values, indicates the relative expression levels of each miRNA. The miRNAs analyzed include miR-335-3p, miR-15b, miR-30d, let-7a, miR-218, miR-141, miR-19b, and miR-200b. Higher ΔCt values correspond to lower expression levels, and vice versa. Symbol indicates * (single star): p<0.05, ** (double star): p<0.01, *** (triple star): p<0.001, **** (four stars): p<0.0001, ns: non-significant.

### 3.2 Genetic and Pedigree Analysis of a Proband

WES identified a homozygous variant in the WNT10A gene, specifically p.Val145Met (c.433G>A) (Figure 3). This variant is classified as “Likely Pathogenic” according to ACMG guidelines and is associated with Tooth Agenesis (OMIM: 606268), following an autosomal recessive (AR) inheritance pattern. The variant is a known mutation.

**Figure 3.**
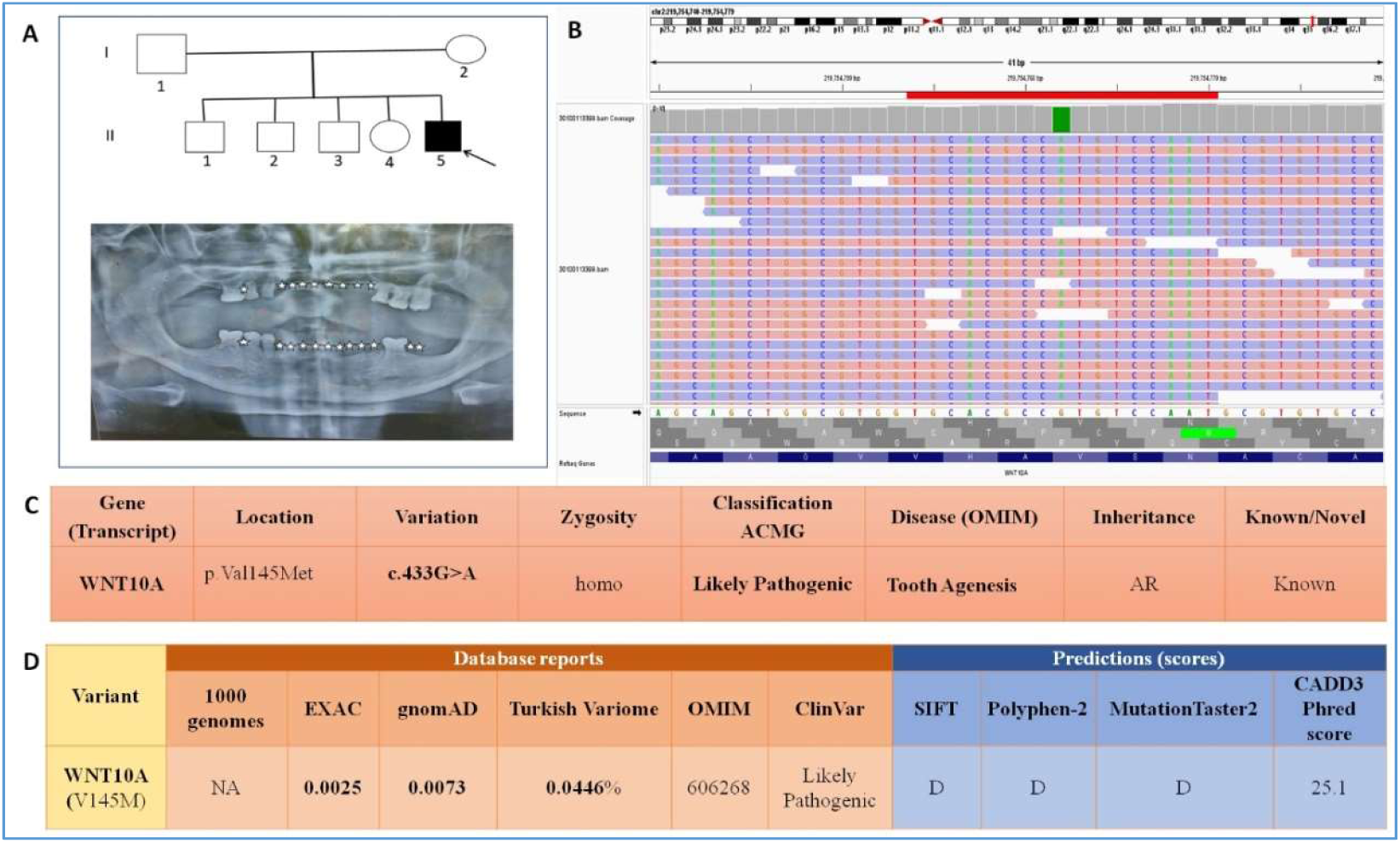
Genetic and Pedigree Analysis of a Proband with Multiple Missing Teeth. **A.** Pedigree analysis identifies the proband with multiple missing teeth, as confirmed by OPG. **B.** Homozygosity mapping reveals the homozygous nature of the identified variants. **C.** Detailed curation of the proband’s genetic profile after WES uncovers a variation in the candidate gene WNT10A, including a known mutation. **D.** The variant’s minor allele frequency (MAF) across different databases and various predictive software analyses indicate its deleterious nature.

Population frequency data show the following minor allele frequencies: 0.0025 in the EXAC database, 0.0073 in gnomAD, and 0.0446% in the Turkish Variome. Functional prediction tools consistently classify this variant as damaging: SIFT(Vaser et al., 2016) predicts it as “Damaging” (D), PolyPhen-2 (Adzhubei et al., 2013) scores it as “Probably Damaging” (D), and MutationTaster2 (Schwarz et al., 2014) also labels it as “Disease Causing” (D). The variant has a CADD Phred score (Rentzsch et al., 2019) of 25.1, indicating a high likelihood of pathogenicity (Figure 3).

### 3.3 Association of miRNA with CTA Gene Expression Data

The association between the gene expression panel and the miRNA target panel revealed that the target genes of miR-218-5p, miR-15b-5p, and miR-200b-3p, which were upregulated, overlapped with six, seven, and three downregulated genes, respectively. These overlapping genes were subsequently analyzed to explore their roles in various pathways and biological functions. Additionally, the downregulated miR-let-7a-3p was found to target 29 genes that coincided with the upregulated genes in the expression panel, and these shared genes were further analyzed for their associated pathways and biological functions (Figure 4).

**Figure 4.**
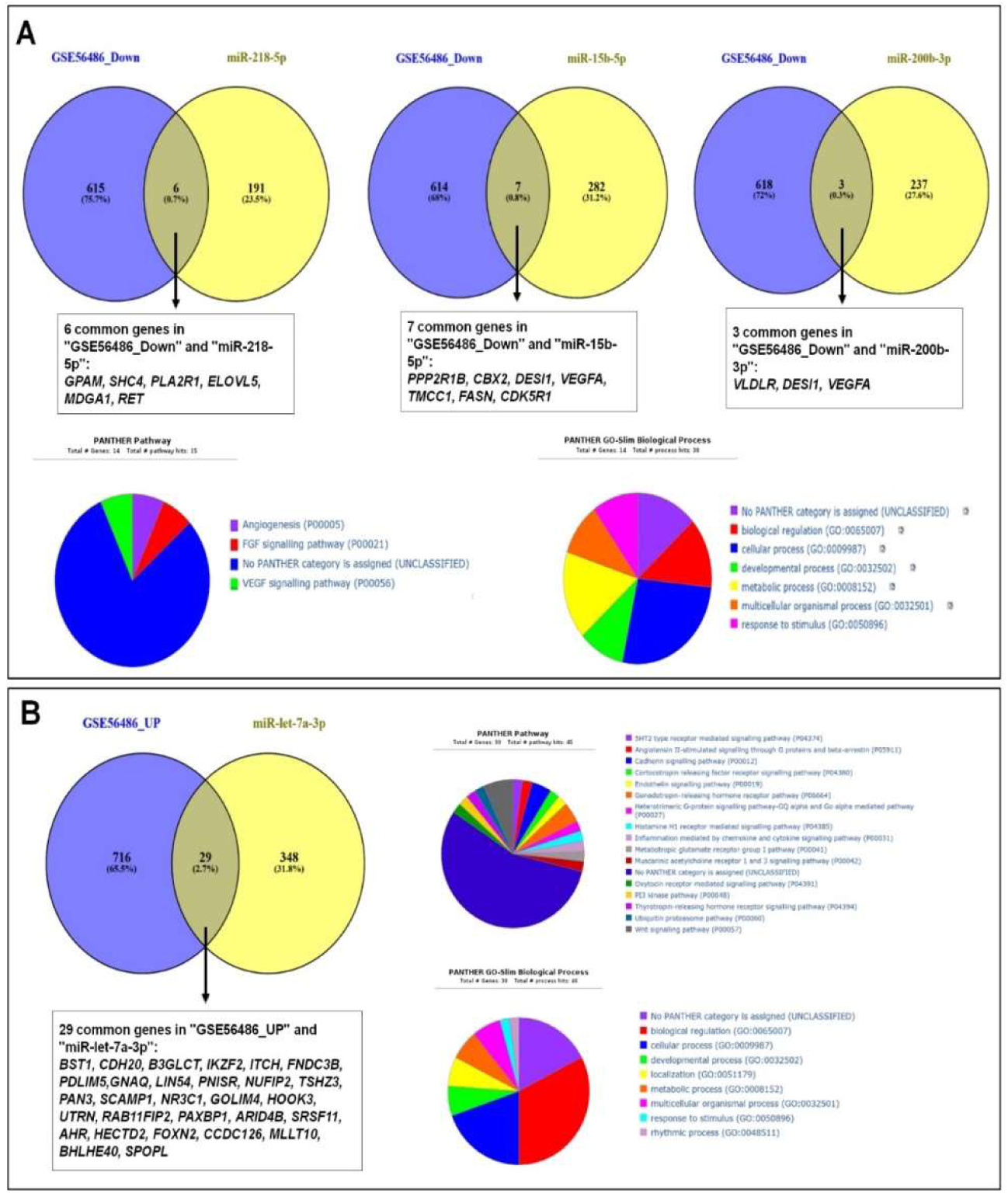
Correlation Analysis of Gene Expression Panels with miRNA Target Panels and Pathway Associations. **A.** Correlation analysis between the gene expression panel and the miRNA target panel reveals that the target genes of upregulated miR-218-5p, miR-15b-5p, and miR-200b-3p overlap with six, seven, and three downregulated genes, respectively. These common genes were further analyzed for their involvement in pathways and biological functions. **B.** The downregulated miR-let-7a-3p targets 29 genes that overlap with the upregulated gene expression panel. These common genes were subsequently analyzed for their associated pathways and biological functions.

### 3.4 Association of miRNA with CTA WES Panel Data

Target genes for the regulated miRNAs were identified through correlation analysis between the WES panel for tooth agenesis and the miRNA target panel. Specifically, 12, 18, 23, and 17 genes were targeted by miR-218-5p, miR-15b-5p, miR-let-7a-3p, and miR-200b-3p, respectively. These overlapping genes were then examined to determine their roles in key biological processes and functions (Figure 5).

**Figure 5.**
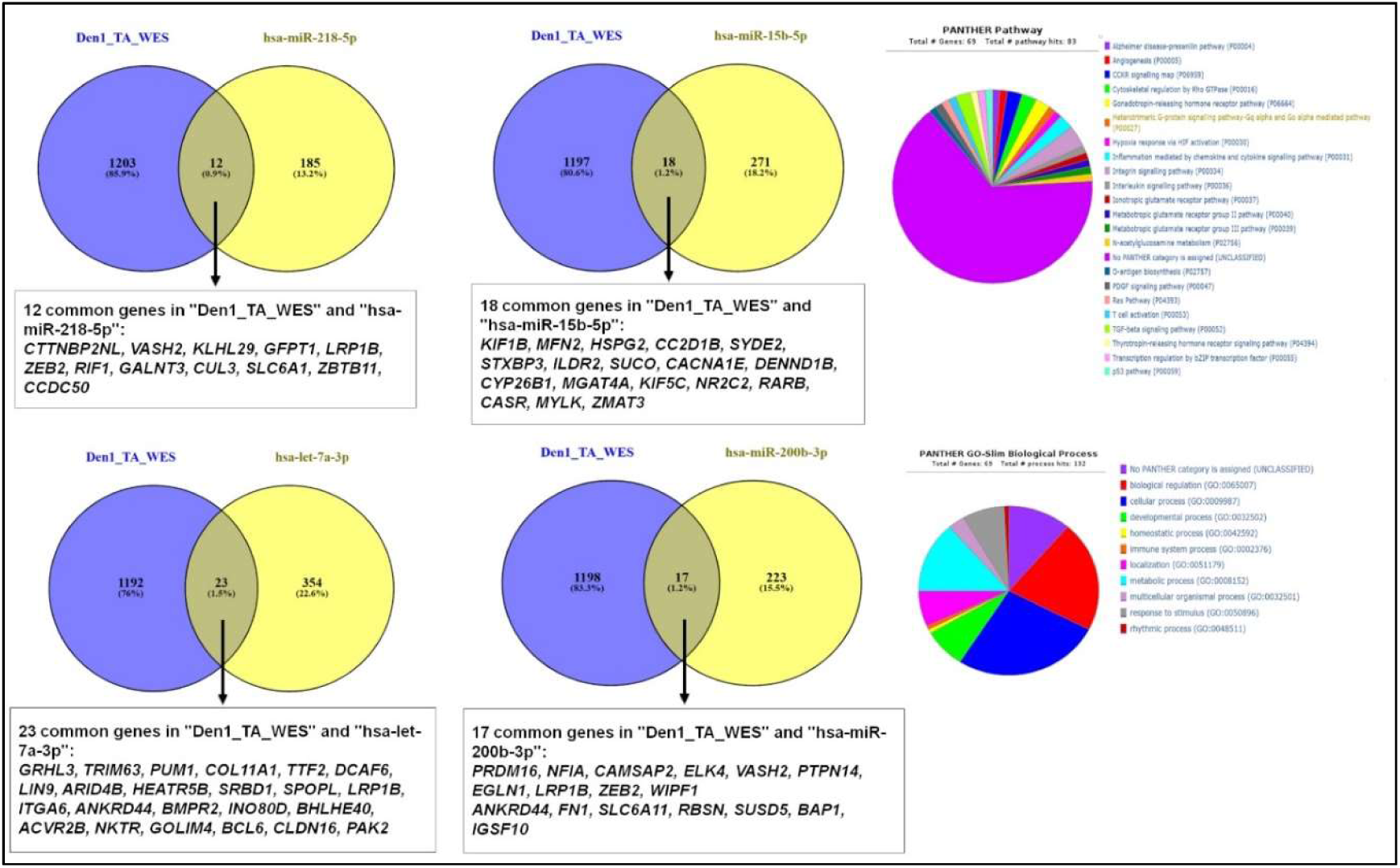
Correlation Analysis of Den1_Tooth Agenesis Panels (WES Panel) with miRNA Target Panels and Pathway Associations. Correlation analysis between the WES Panel for tooth agenesis (Den1) and the miRNA target panel reveals overlapping target genes for the regulated miRNAs. Specifically, miR-218-5p targets 12 genes, miR-15b-5p targets 18 genes, miR-let-7a-3p targets 23 genes, and miR-200b-3p targets 17 genes. These common genes were further investigated for their involvement in relevant pathways and biological functions.

### 3.5 Multi-omics Approaches

In the multi-omics analysis of miR-218-5p, 22 common elements were identified across different panels (Figure 6-7). Specifically, five common genes (*GPR161, SV2A, GNB1, GPR153, IKBKB*) were shared between the miR-218-5p target panel and the Metabolite Panel.

**Figure 6.**
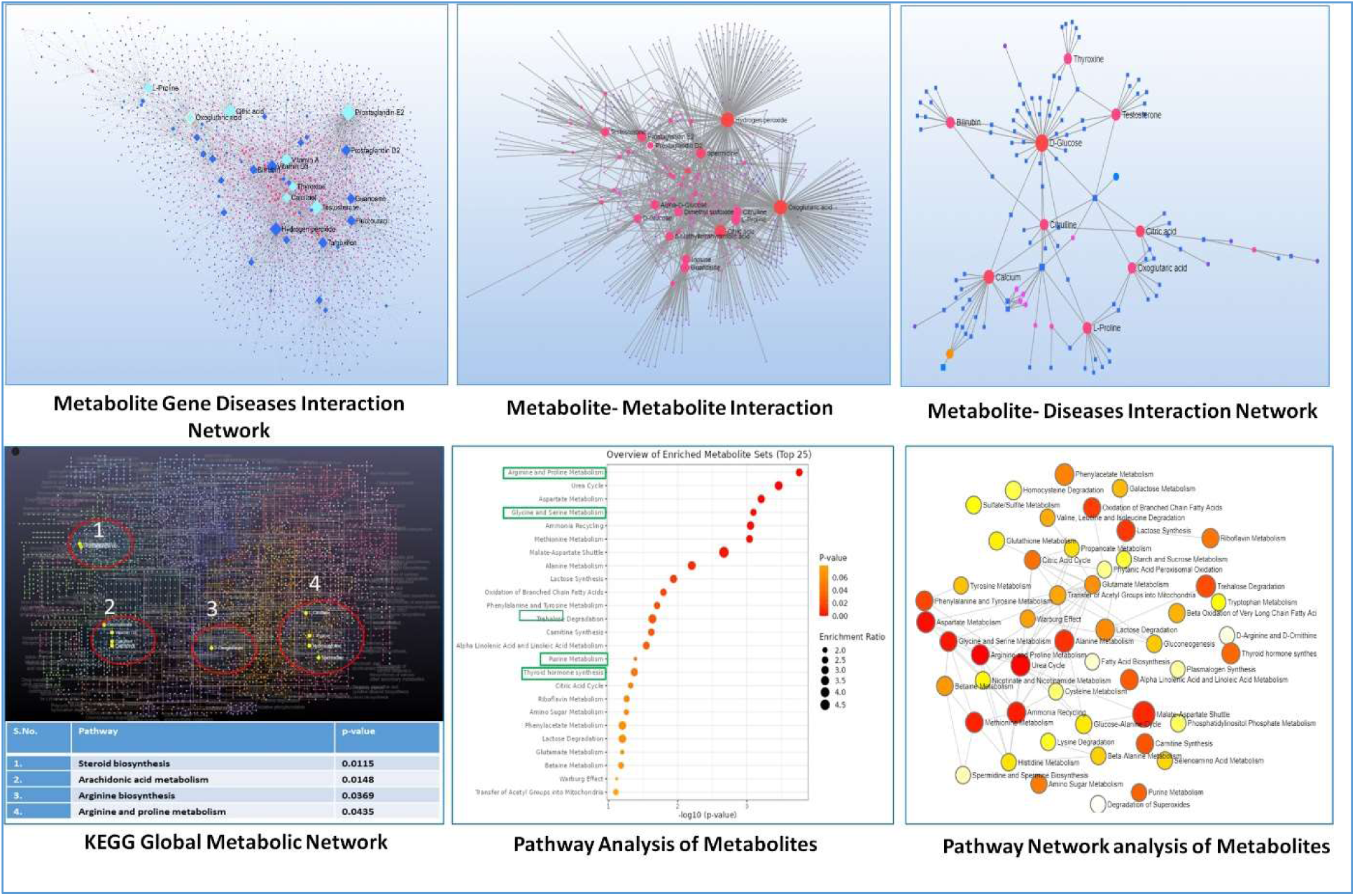
Role of Metabolites in Tooth Agenesis: Interaction Networks and Pathway Analysis. The Metabolite-Gene-Disease Interaction Network illustrates how metabolites are connected to various diseases. Metabolites associated with tooth development diseases are highlighted in sky blue, while smaller nodes, appearing as dots, represent other diseases. The Metabolite-Metabolite Interaction Network shows how a hub metabolite interacts with multiple other metabolites. The Metabolite-Disease Interaction Network further reveals direct associations between specific metabolites and various diseases. The KEGG global metabolite network analysis identifies four pathways involved in the CTA process. Pathway analysis highlights several pathways directly involved in tooth development, which are enclosed in green rectangles. The network visualizes these pathway interactions, with node size representing enrichment scores and node color indicating p-value significance, where red and blue reflect the range of significance levels.

**Figure 7.**
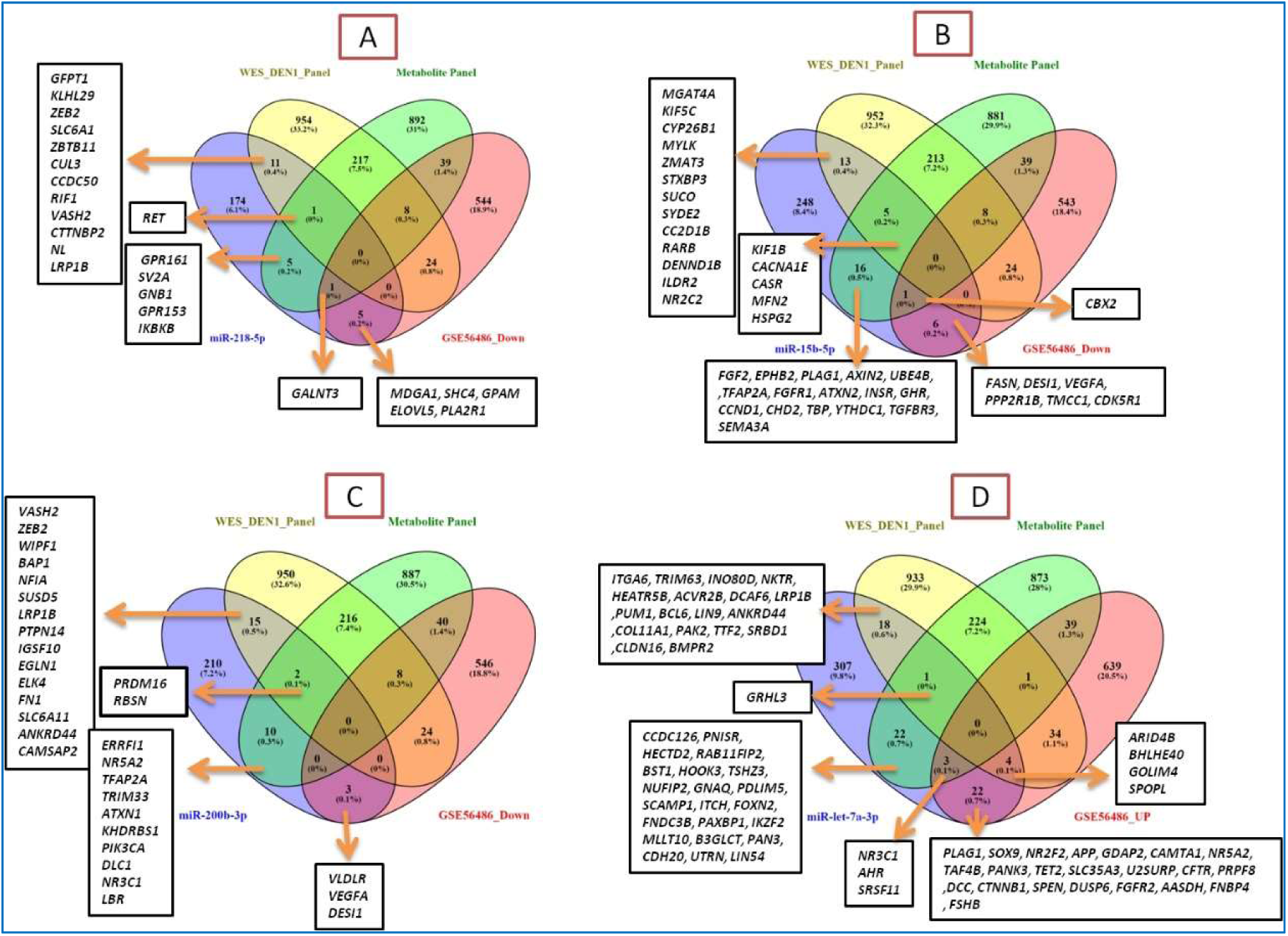
Cumulative Analysis of Four miRNA Target Panels with WES, Metabolite, and Gene Expression Panels. **A, B, C,** and **D** represent the cumulative analysis of the target panels for miR-218-5p, miR-15b-5p, miR-200b-3p, and miR-let-7a-3p, respectively, in relation to the WES, metabolite, and gene expression panels. These analyses reveal the overlapping genes shared between the different panels.

The RET gene was identified as common across the miR-218-5p target panel, the Metabolite Panel, and the GSE56486 Down panel. Additionally, five genes (*MDGA1, SHC4, GPAM, ELOVL5, PLA2R1*) were found to overlap between the miR-218-5p target panel and the GSE56486 Down panel. The gene *GALNT3* was common to the miR-218-5p target panel, the WES DEN1 Panel, and the Metabolite Panel. Lastly, 11 common genes (*GFPT1, KLHL29, ZEB2, SLC6A1, ZBTB11, CUL3, CCDC50, RIF1, VASH2, CTTNBP2NL, LRP1B*) were found between the miR-218-5p target panel and the WES DEN1 Panel (Figure 7).

This comprehensive integration of miR-218-5p with different gene panels underscores the relevance of these overlapping genes in the context of miRNA-mediated regulation in tooth development and associated pathways.

miR-15b-5p demonstrated notable overlap with various gene panels in the multi-omics study. Thirteen genes, specifically, shared characteristics between the WES DEN1 Panel and the miR-15b-5p target panel: *MGAT4A, KIF5C, CYP26B1, MYLK, ZMAT3, STXBP3, SUCO, SYDE2, CC2D1B, RARB, DENND1B, ILDR2, NR2C2*. Additionally, the *CBX2* gene was identified across the miR-15b-5p target panel, the Metabolite Panel, and the GSE56486 Down panel (Figure 7). Five genes (*KIF1B, CACNA1E, CASR, MFN2, HSPG2*) were shared among the miR-15b-5p target panel, the WES DEN1 Panel, and the Metabolite Panel. Moreover, 16 genes (*FGF2, EPHB2, PLAG1, AXIN2, UBE4B, TFAP2A, FGFR1, ATXN2, INSR, GHR, CCND1, CHD2, TBP, YTHDC1, TGFBR3, SEMA3A*) were common between the miR-15b-5p target panel and the Metabolite Panel (Figure 7). Finally, six genes (*FASN, DESI1, VEGFA, PPP2R1B, TMCC1, CDK5R1*) overlapped between the miR-15b-5p target panel and the GSE56486 Down panel. These overlapping genes underscore the potential regulatory role of miR-15b-5p in tooth development and related pathways (Figure 7-9).

Our investigation of miR-200b-3p identified numerous important relationships. Three common elements (*VLDLR, VEGFA,* and *DESI1*) were identified between miR-200b-3p and the GSE56486 Down panel, highlighting their involvement in downregulated pathways. Additionally, two common elements (*PRDM16* and *RBSN*) were found between miR-200b-3p, the WES DEN1 Panel, and the Metabolite Panel, indicating their potential regulatory role across these datasets (Figure 7). Furthermore, 15 common elements (*VASH2, ZEB2, WIPF1, BAP1, NFIA, SUSD5, LRP1B, PTPN14, IGSF10, EGLN1, ELK4, FN1, SLC6A11, ANKRD44, CAMSAP2*) were shared between miR-200b-3p and the WES DEN1 Panel, suggesting a strong link between miR-200b-3p and the genetic basis of tooth agenesis (Figure 7). Finally, 10 common elements (*ERRFI1, NR5A2, TFAP2A, TRIM33, ATXN1, KHDRBS1, PIK3CA, DLC1, NR3C1, LBR*) were identified between miR-200b-3p and the Metabolite Panel, underscoring the potential metabolic implications of miR-200b-3p in tooth development.

The analysis of miR-let-7a-3p shows significant overlap with genes across various datasets related to tooth development. Specifically, three genes (*NR3C1, AHR*, and *SRSF11*) were identified as common between miR-let-7a-3p, the Metabolite Panel, and the GSE56486 UP panel. Additionally, four common genes (*ARID4B, BHLHE40, GOLIM4*, and *SPOPL*) were shared between miR-let-7a-3p, the WES DEN1 Panel, and GSE56486 UP (Figure 7).. Further analysis revealed 22 common genes between the miR-let-7a-3p target panel and the WES DEN1 panel. These genes, which include *BMP2, CALM1, CCND1, CDK5, COL5A2, FGF2, FOXP1, FN1, FOXO3, HOXA9, NOTCH2, NR2F2, PRDM16, PRKCB, RUNX2, SHC1, SOS1, SOX4, TCF7L2, TGFBR2, ZEB2*, and *ZEB1*, underline the strong genetic associations with miR-let-7a-3p in tooth development.

### 3.6 Biological and pathway analysis

In our analysis, the target panels of miR-218-5p, miR-15b-5p, and miR-200b-3p, in comparison to WES and gene expression panels, revealed that tooth development is driven by key biological processes, including developmental processes (GO:0032502), biological regulation (GO:0065007), cellular processes (GO:0009987), and metabolic processes (GO:0008152). These processes collectively support tissue formation, differentiation, and the intricate regulation necessary for proper tooth morphogenesis (Figure 8-9).

**Figure 8:**
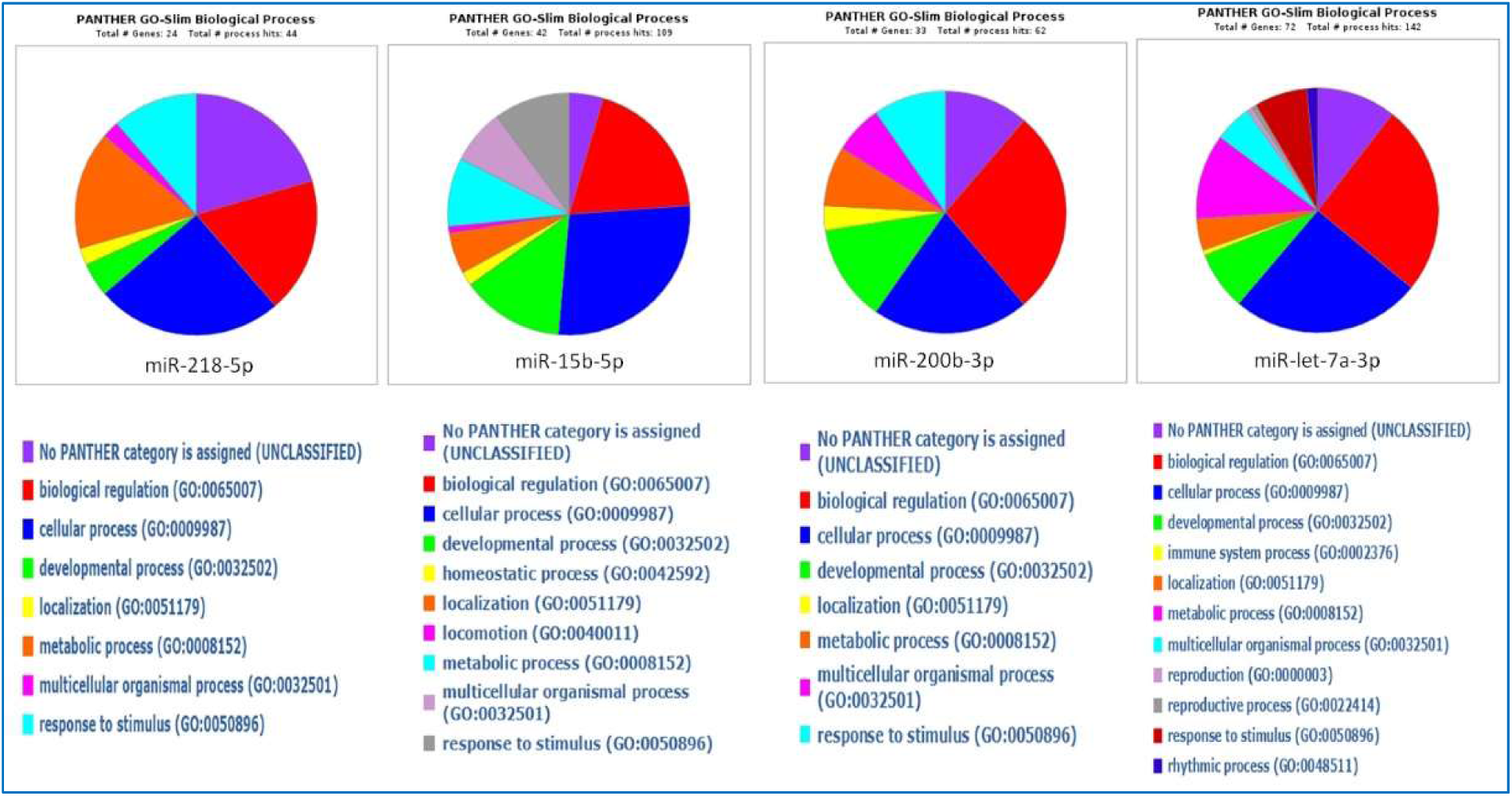
Biological Functions of Overlapping Genes from Panel Analyses of Four Individual miRNAs. This figure illustrates the biological functions of all genes that overlap across the different panel analyses (WES, metabolite, and gene expression) for each of the four miRNAs: miR-218-5p, miR-15b-5p, miR-200b-3p, and miR-let-7a-3p.

**Figure 9:**
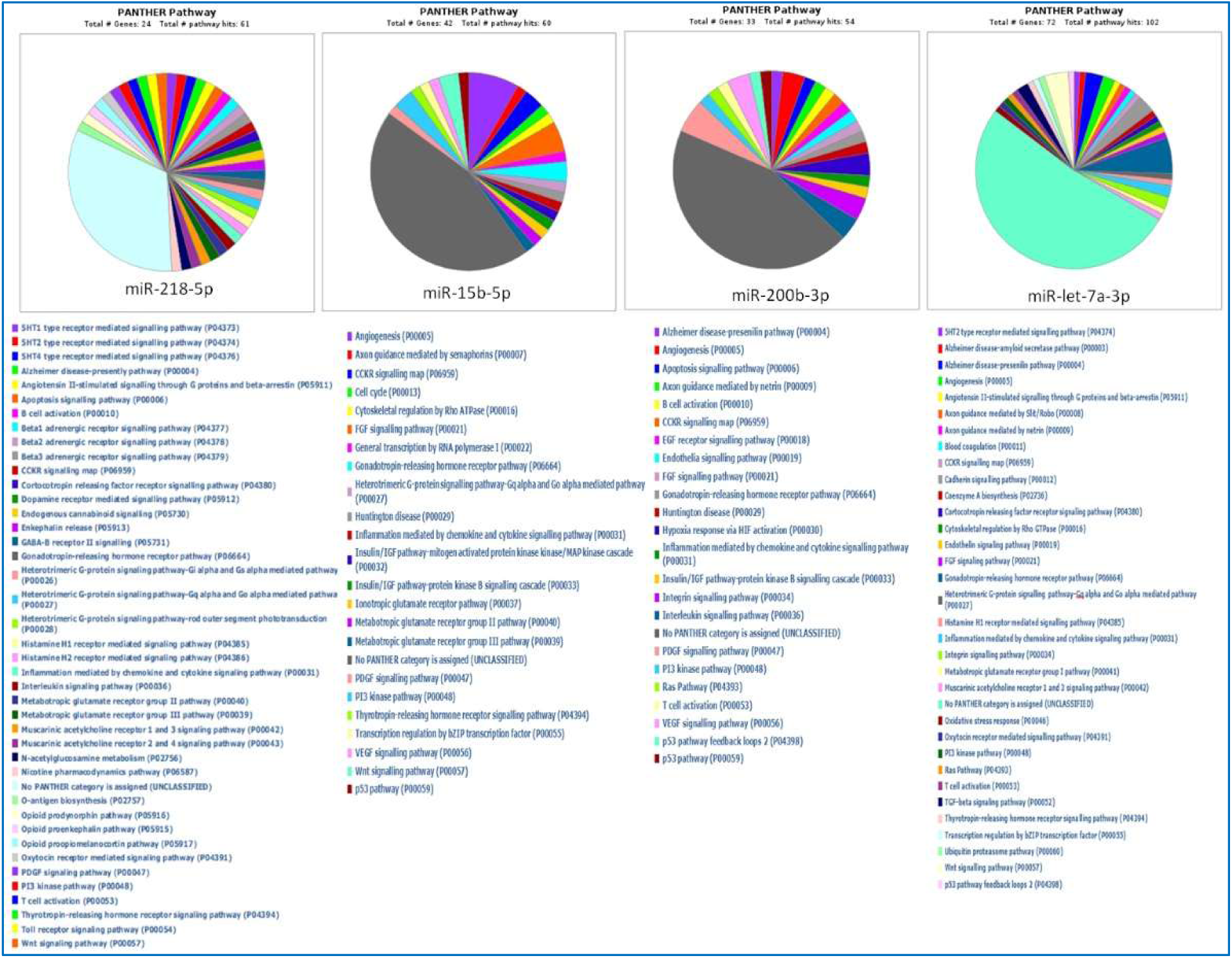
Pathway Analysis of Overlapping Genes from Panel Analyses of Four Individual miRNAs. This figure presents the pathway analysis of genes that overlap across the different panel analyses (WES, metabolite, and gene expression) for each of the four miRNAs: miR-218-5p, miR-15b-5p, miR-200b-3p, and miR-let-7a-3p.

Pathway analysis highlighted the Integrin signaling (P00034) and PDGF signaling (P00047) as central to the formation and patterning of dental tissues. For miR-let-7a-3p, the analysis underscored the importance of the TGF-beta signaling pathway (P00052) alongside the Integrin signaling pathway. In the gene expression panel, additional pathways such as angiogenesis, FGF, and VEGF signaling were identified as crucial for vascular support, early cell growth, and differentiation necessary for tooth maturation. The Wnt, PI3 kinase, and Cadherin signaling pathways, along with inflammation and ubiquitin-proteasome pathways, were noted for their roles in cellular growth, adhesion, immune modulation, and protein regulation during tooth development (Figure 8-9).

The cumulative results of these investigations highlight the crucial role played by these miRNAs in regulating the intricate biological processes and pathways necessary for tooth growth.

The Tri-panel Analysis of the four miRNAs (miR-218-5p, miR-15b-5p, miR-200b-3p, and let-7a-3p) has provided significant insights into the biological functions and pathways involved in tooth development. The analysis of the four miRNAs (miR-218-5p, miR-15b-5p, miR-200b-3p, and let-7a-3p) revealed common biological functions and pathways that are crucial for tooth development. The shared biological functions include biological regulation (GO:0065007), cellular processes (GO:0009987), developmental processes (GO:0032502), localization (GO:0051179), metabolic processes (GO:0008152), and multicellular organismal processes (GO:0032501). These functions collectively contribute to the regulation of gene expression, coordination of cellular activities, tissue organization, and the metabolic and developmental processes essential for the formation and maintenance of dental tissues (Figure 8-9).

Pathway analysis identified several common pathways among these miRNAs that are instrumental in tooth development. The Wnt signaling pathway (P00057) plays a key role in regulating cell proliferation and differentiation necessary for tooth formation. Angiogenesis (P00005) is crucial for forming blood vessels that supply nutrients to developing teeth. The FGF signaling pathway (P00021) influences early tooth development through cell growth and tissue organization, while the PI3 kinase pathway (P00048) promotes cell survival and growth. Additionally, inflammation mediated by chemokine and cytokine signaling (P00031) modulates the immune response, ensuring a healthy environment for tooth development (Figure 8-9).

These findings underscore the significance of these miRNAs in orchestrating the biological functions and pathways essential for proper tooth development.

### 3.7 Network Analysis and Identification of Hub Genes Targeted by MiRNAs

To explore the potential regulatory roles of the selected miRNAs in tooth development, we performed a network analysis focusing on their target genes. This analysis identified several key hub genes (Figure 10) that are likely pivotal in the associated pathways. miR-let-7a-3p was found to target several significant genes, including *CTNNB1, SOX9, GNAQ, ITGA6, APP, COL18A1,* and *BMPR2*. For miR-15b-5p, the analysis revealed connections with key genes such as *FGF2, CCND1, FGFR2, FGRF1, AXIN2, HSPG2, SEMA3A, CASR,* and *TBP*. miR-200b-3p was associated with the genes *FN1, PIK3CA,* and *ZEB2*. miR-218-5p was also found to target *ZEB2* in addition to *RET*. The involvement of *ZEB2* as a common target for both miR-200b-3p and miR-218-5p highlights its potential as a critical hub gene in the regulatory network.

**Figure 10:**
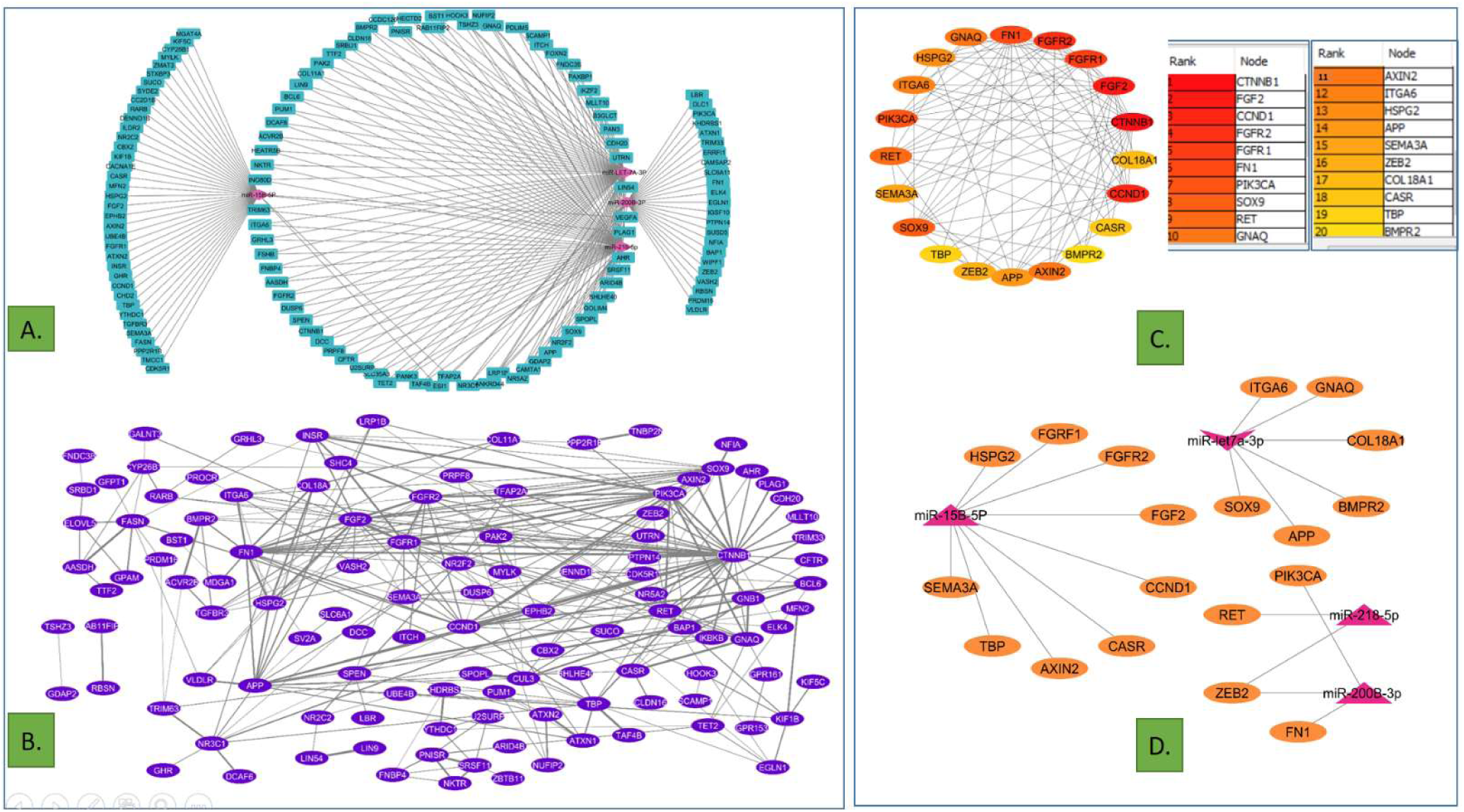
Identification of Hub Genes through Network Analysis of miRNA-Targeted Shared Genes from Multi-omics Panels. **A.** Network analysis of miRNAs and their shared gene targets, highlighting the interactions within the multi-omics-derived dataset. **B.** Network analysis of shared genes using STRING to explore the connections between these genes. **C.** The top 20 hub genes identified from the analysis in panel B, ranked based on the MCC method using the CytoHubba plugin. **D.** Network visualization showing the specific miRNAs targeting the identified hub genes, demonstrating the distinct regulatory influence of different miRNAs.

## 4. Discussion

Our study offers critical insights into the regulatory role of miRNAs in tooth development, with a particular emphasis on congenital tooth agenesis (CTA). By employing a multi-omics strategy, we identified four key miRNAs—miR-218-5p, miR-15b-5p, miR-200b-3p, and miR-let-7a-3p—that are significantly associated with CTA. This was confirmed through RT-qPCR analysis and validated across WES, metabolite, and gene expression panels.

The novelty of our research lies in the systematic identification and validation of these specific miRNAs as pivotal regulators in tooth development, particularly in the context of CTA. Notably, miR-218-5p and miR-let-7a-3p stand out for their previously unrecognized roles in dental tissue morphogenesis, with miR-218-5p being significantly involved in critical, yet previously unidentified, signaling networks essential for tooth development.

### 4.1 miR-218-5p: A Key Regulator in Tooth Development

The study identifies miR-218-5p as a novel miRNA in the context of tooth development, with no prior associations documented in dental research. Through multi-omics analysis, miR-218-5p is shown to target key genes across multiple biological pathways, suggesting its pivotal role in regulating odontogenesis. Specifically, the multi-omics approach revealed the novel involvement of miR-218-5p in targeting RET, a gene crucial for craniofacial and dental development, along with *ZEB2*, a regulator of epithelial-to-mesenchymal transition (EMT) (Lamouille et al., 2014). The downregulation of *ZEB2*, due to the elevated levels of miR-218-5p, could disrupt the normal EMT process, potentially leading to tooth agenesis by impairing the proper formation of dental tissues.

The identification of RET as a common element across multiple panels (the miR-218-5p target panel, Metabolite Panel, and GSE56486 Down panel) underscores its significant, novel role in tooth development mediated by miR-218-5p. RET mutations have been associated with various developmental disorders, including Hirschsprung disease, which also involves craniofacial abnormalities (Airaksinen & Saarma, 2002). Therefore, miR-218-5p’s regulation of RET further highlights its pivotal role in maintaining the integrity of the signaling networks necessary for proper odontogenesis.

### 4.2 miR-15b-5p: Influencing Growth Factor Signaling in Odontogenesis

miR-15b-5p, another miRNA upregulated in CTA, was found to target a broad array of genes, many of which are involved in growth factor signaling pathways. Notably, miR-15b-5p targets *FGF2, CCND1*, and *FGFR2*, all of which are critical components of the fibroblast growth factor (*FGF*) signaling pathway (Thesleff & Sharpe, 1997).

The FGF signaling pathway is indispensable for tooth development, particularly during the early stages of odontogenesis where it regulates the proliferation and differentiation of dental epithelial and mesenchymal cells (Nie et al., 2006). Dysregulation of this pathway, due to aberrant expression of miR-15b-5p, can lead to developmental defects such as hypodontia or oligodontia, forms of CTA characterized by the congenital absence of multiple teeth. FGFR2 mutations, in particular, have been associated with syndromic forms of tooth agenesis, further underscoring the importance of this signaling axis in dental development.

Moreover, the overlap of miR-15b-5p targets with genes involved in key metabolic pathways, as revealed by the multi-omics analysis, suggests that miR-15b-5p might also influence the metabolic environment essential for tooth development (Balmer & Blomhoff, 2002). Metabolites such as amino acids and lipids, regulated by genes in the miR-15b-5p target network, provide the necessary building blocks for the synthesis of dental tissues, thereby implicating miR-15b-5p in both the growth signaling and metabolic regulation of odontogenesis.

### 4.3 miR-200b-3p: Linking EMT and Extracellular Matrix Remodeling

miR-200b-3p was identified as significantly upregulated in CTA and has been associated with the regulation of genes involved in EMT and extracellular matrix (ECM) remodeling, two processes critical for tooth development. Among its key targets, miR-200b-3p regulates *ZEB2* and *FN1* (fibronectin 1), both of which play vital roles in the structural organization of dental tissues (Gheldof & Berx, 2013).

*ZEB2*, as discussed earlier, is a major regulator of EMT, and its inhibition by miR-200b-3p could interfere with the migration and differentiation of dental progenitor cells. The ability of miR-200b-3p to target both EMT regulators and ECM components such as FN1 suggests that it plays a dual role in coordinating the cellular and structural aspects of tooth formation.

Furthermore, miR-200b-3p’s involvement in ECM remodeling is critical for the deposition and organization of dental matrices, such as dentin and enamel. The ECM provides the scaffold necessary for mineralization, and disruptions in ECM homeostasis can lead to dental abnormalities, including hypomineralization and tooth agenesis(Butcher et al., 2009). The overlap of miR-200b-3p targets with genes involved in ECM and EMT pathways highlights its comprehensive role in orchestrating the complex interactions required for normal tooth development.

### 4.4 miR-let-7a-3p: A Potential Inhibitor of Tooth Development Pathways

While miR-let-7a-3p has been previously associated with various biological processes, its specific downregulation in CTA, and the subsequent impact on tooth development, is a novel finding of this research. The multi-omics analysis highlighted *CTNNB1* (β-catenin) as a novel target gene associated with miR-let-7a-3p, linking it to the Wnt signaling pathway, which is essential for tooth formation (Iaquinta et al., 2021). A reduction in miR-let-7a-3p could lead to the overactivation of Wnt signaling, potentially causing aberrant tooth development and contributing to the pathogenesis of CTA. (Bell et al., 1997). The downregulation of miR-let-7a-3p may therefore contribute to an imbalance in these signaling networks, promoting conditions conducive to tooth agenesis.

In our study, the tri-panel analysis of miR-218-5p, miR-15b-5p, miR-200b-3p, and let-7a-3p revealed critical biological processes and pathways integral to tooth development. Key processes include biological regulation, cellular and developmental processes, and metabolic activities, all of which support tissue differentiation and morphogenesis. Pathway analysis highlighted the roles of *Wnt*, Integrin, *PDGF*, *TGF*-beta, and *FGF* signaling, alongside angiogenesis and *PI3* kinase pathways, underscoring their contributions to cell growth, differentiation, and the vascular support necessary for proper tooth formation. These findings affirm the miRNAs’ role in regulating essential pathways for dental tissue development.

### 4.5 Integrating miRNA and Genetic Data: Implications for CTA Pathogenesis

The integration of miRNA expression data with WES and metabolomic profiles in this study provides a comprehensive view of the molecular mechanisms underlying CTA. The overlaps between miRNA target genes and those identified in WES and metabolomic analyses point to a complex interplay between genetic predisposition and miRNA-mediated regulatory networks in the etiology of CTA.

The identification of the hub genes through multi-omics analysis highlights the complex regulatory networks governing tooth development and the role of specific miRNAs in this process. miRNAs like miR-let-7a-3p, miR-15b-5p, miR-200b-3p, and miR-218-5p have emerged as critical regulators by targeting genes that are integral to signaling pathways and cellular processes involved in odontogenesis.

For example, the identified homozygous variant in the *WNT10A* gene, known to be associated with tooth agenesis, may interact with the regulatory networks governed by miRNAs such as miR-218-5p and miR-200b-3p, both of which target components of the Wnt signaling pathway. This interaction could exacerbate the effects of genetic mutations, leading to a more pronounced phenotype.

Moreover, the multi-omics approach highlighted the role of metabolic pathways in tooth development, with miRNAs such as miR-15b-5p and miR-let-7a-3p regulating genes involved in key metabolic processes. These findings suggest that metabolic dysregulation, in conjunction with genetic and epigenetic factors, may contribute to the development of CTA.

## 5. Conclusion

This study provides novel insights into the role of specific miRNAs in CTA, highlighting their potential as biomarkers and therapeutic targets. Through comprehensive bioinformatics analysis and multi-omics integration, we identified four key miRNAs—miR-218-5p, miR-15b-5p, miR-200b-3p, and let-7a-3p—that are significantly associated with CTA and tooth development. The multi-omics approach, incorporating gene expression data, metabolite analysis, and WES panels, supported the involvement of these miRNAs in critical biological pathways, such as the *Wnt* signaling, *FGF* signaling, and *PI3* kinase pathways, which are essential for proper tooth formation.

The identification of these miRNAs in blood samples, rather than traditional dental tissues, represents a significant advancement in the non-invasive diagnosis and management of dental anomalies. Our findings suggest that these miRNAs play crucial roles in biological regulation, cellular processes, and developmental pathways, emphasizing their potential as biomarkers for early diagnosis and personalized treatment strategies.

Overall, this study contributes to the growing understanding of miRNA-mediated regulation in odontogenesis and demonstrates the value of multi-omics approaches in uncovering complex gene networks. The findings open new avenues for research into therapeutic approaches targeting miRNAs to treat or prevent congenital dental anomalies. Further investigation is warranted to explore the clinical application of these findings and to fully elucidate the miRNA-mRNA networks involved in tooth development and related disorders.

## Supporting information

Supplementaryfile2_genepanel

## Data Availability

All data produced in the present study are available upon reasonable request to the authors

## Abbreviations

CTA: Congenital Tooth Agenesis
WES: Whole Exome Sequencing
DGE: Differential Gene Expression
EOC: Epithelial Ovarian Cancer
HCC: Hepatocellular Carcinoma
OSCC: Oral Squamous Cell Carcinoma
OC: Ovarian Cancer
B-CLL: B-cell Chronic Lymphocytic Leukemia
APL: Acute Promyelocytic Leukemia
CC: Colorectal Cancer
ASD: Autism Spectrum Disorder
MM: Multiple Myeloma
HNSCC: Head and Neck Squamous Cell Carcinoma
PDAC: Pancreatic Ductal Adenocarcinoma
DM: Dermatomyositis
DMD: Duchenne Muscular Dystrophy
FSHD: Facioscapulohumeral Muscular Dystrophy
LGMD2A: Limb-Girdle Muscular Dystrophies Types 2A
NM: Nemaline Myopathy
ALL: Acute Lymphoblastic Leukemia
PBC: Primary Biliary Cirrhosis
CLL: Chronic Lymphocytic Leukemia
ATC: Anaplastic Thyroid Carcinoma
NAFLD: Non-Alcoholic Fatty Liver Disease

## 6. Acknowledgement

We gratefully acknowledge the Indian Council of Medical Research (ICMR) for providing the Senior Research Fellowship (SRF) to PR and CD. We extend our sincere thanks to Dr. Garima Jain, MPDF, CGD, Banaras Hindu University (BHU), Varanasi, for her valuable support and insightful contributions to our research on miRNAs.

## 7. Author Contribution

**PR** conceived the research idea, conducted experiments, performed bioinformatics and data analysis, and wrote the manuscript. **CD** contributed to data analysis. **NV** was responsible for identifying CTA patients and conducting OPG analysis. **RB** and **VKS** carried out clinical investigations. **PD** co-conceived the research idea and approval of final draft.

## 8. Declaration

The authors declare no conflict of interest.

## Notes

### Competing Interest Statement

The authors have declared no competing interest.

### Funding Statement

This study did not receive any funding

### Author Declarations

Institute Ethics committee, Institute of Science, BHU, Varanasi

