## Supplementary file 1 for "Understanding Tooth Agenesis: A Multi-omics Insight into MicroRNA Regulation"

**Table1: miRNA Regulation in Dental and Cancerous Cells**

| **Cell Type** | **miRNAs** | **Possible Functions** | **References** |
| --- | --- | --- | --- |
| Ameloblasts | miR-224, miR-29b | Enamel mineralization | Fan et al (2015b), Chen et al. (2015) |
| Alveolar bone cells | miR-101, miR-31, miR-146a | Tooth movement | Chen et al (2014), Ge et al. (2015), Klingelhoffer et al (2016) |
| Dental follicle cells | miR-34a | Differentiation of dental follicle cells, tooth eruption | Wan et al (2012) |
| Dental papilla cells | miR-34a | Differentiation of dental papilla cells | Wan et al (2012) |
| Dental pulp cells | miR-135, miR-135b, miR-141, miR-135b, miR-143, miR-146a, miR-150-3p, miR-155, miR-200a, miR-218, miR-32, miR-34a, miR-424, miR-433, miR-516a-3p, miR- 584, miR-586, miR-7- 5p, miR-720, miR-766, miR-885-5p | Dental pulp repair and regeneration, Differentiation of dental pulp cell, Inflammatory and immunological response, Myogenic differentiation Neuronal development, Odontoblast differentiation, Neuronal development, Odontoblast differentiation | Zhong et al (2012), Li et at (2015), Song et al. (2017), Wang et al. (2015), Liu et al. (2014b), Song et al. (2017), Huang et al (2011), Gay et al. (2014), Hara et al (2013), Sipert et at (2014), Vasanthan et al (2015), Kong et at (2014), Chen et al, (2012) |
| Periodontal ligament cells | miR-30a, miR-379, miR-520d, miR-548a miR-1297, miR-138, miR-141, miR-145-5p, miR-146a, miR-155, miR-182, miR-195, miR-21, miR-24, miR- 200a, miR-218, miR- 224-5p, miR-29b, miR- 3607-5p, miR-424-5p, miR-4328, miR-720 | Bone formation, Cell survival, Differentiation, Inflammatory and immunological response, Tooth movement | Zhou et al. (2016), Chen et al (2016), Chen et al (2015), Gay et al (2014), Hara et al (2013), Liu et al (2022), Du et al (2016), Wang et al (2016), Chang et al (2015), Sipert et al. (2014), Sipert et al. (2014b) |
| Gingival fibroblasts | miR-141, miR-146a, miR-155, miR-200a, miR-218, miR-29b, miR451-5p, miR-223, miR-486, miR-3917, miR-1246, miR-1260, miR-141, miR1260b, miR-203, miR-210, miR-205-3p, miR-223 Let-7a, let-7c, miR- 19b, miR-103, miR- 130a, miR-146a, miR- 15a, miR-150, miR- 181b, miR-199a-5p, miR-23a, miR-200b, miR-214, miR-22, miR-223, miR-30d, miR-301a, miR-106b, miR-130a, miR-142- 3p, miR-185, miR-210 | Inflammatory and immunological response, Tissue homeostasis | Chen et al (2015), Gay et al. (2014), Sipert et al (2014a), Sipert et al. (2014b) |
| Gingival tissue | miR-141, miR-146a, miR-155, miR-200a, miR-218, miR-29b, miR451-5p, miR-223, miR-486, miR-3917, miR-1246, miR-1260, miR-141, miR1260b, miR-203, miR-210, miR-205-3p, miR-223 Let-7a, let-7c, miR- 19b, miR-103, miR- 130a, miR-146a, miR- 15a, miR-150, miR- 181b, miR-199a-5p, miR-23a, miR-200b, miR-214, miR-22, miR-223, miR-30d, miR-301a, miR-106b, miR-130a, miR-142- 3p, miR-185, miR-210 | Inflammatory and immunological response, Tissue homeostasis | Xie et al (2011), Stoecklin- Wasmer et al (2012), Lee et al (2013), Perri et al. (2012), Motedayyen et al. (2015), Ogata et al. (2014) |
|  | miR-9, miR-15b, miR-27, miR-29a, miR-30a, miR-103/107, miR-155, miR-194, miR-200 family, miR-205, miR-204, miR-221/222, miR-661 | miRNAs involved in Epithelial Mesenchymal Transition and Mesenchymal Epithelial Transition (EMT/MET) | Zhang et al. 2012 |
|  | miR-let7, miR-7, miR-10a, miR-10b, miR-16, miR-17-92, miR-21, miR-22, miR-31, miR-122, miR-126, miR-146a/b, miR-194, miR-206, miR-214, miR-335 | Additional miRNA (besides those regulating EMT/MET) with functional roles in tumor invasion and metastasis | Dang et al. 2014 |
| Dental epithelium | miR-373, miR-378, miR-520c | Tooth evolution and dental defects | Rutanam et al.2013 |
| Dental mesenchyme | miR-31, miR-140, miR-141, miR-455, miR-689, miR-711, 720, miR-875-5p, miR- 200 family | Tooth evolution and dental defects | Sehic et al.2017, Kuhu et al.2016 |
| Dental epithelium and mesenchyme | miR-689, miR-711, miR-720, miR-183 | Tooth evolution and dental defects | Michon et al. 2010 |

**Table2: Overlapping Targets Between Selected miRNAs and the Congenital Tooth Agenesis (CTA) Gene Panel.**

| miRNA | **Functional work** | **Targets** | **No of Targets** |
| --- | --- | --- | --- |
| hsa-miR-335-3p | Functional roles in tumor invasion and metastasis | LRP2 | 9 |
|  |  | PGAP1 |  |
|  |  | ITGB6 |  |
|  |  | HOXD13 |  |
|  |  | PIGV |  |
|  |  | TMEM237 |  |
|  |  | PAX8 |  |
|  |  | IGSF3 |  |
|  |  | DPYD |  |
| hsa-miR-424-5p | Bone formation, Cell Survival, Differentitation Inflammatory and immunological response, Tooth movement | KIF1B | 7 |
|  |  | AKT3 |  |
|  |  | MFN2 |  |
|  |  | HSPG2 |  |
| hsa-miR-15b-5p | Involved in Epithelial Mesenchymal Transition and Mesenchymal Epithelial Transition | KIF1B | 5 |
|  |  | AKT3 |  |
|  |  | MFN2 |  |
|  |  | WNT3A |  |
|  |  | HSPG2 |  |
| hsa-miR-200c-3p | Tooth development, Dental pulp repair and regeneration, Differentiation of dental pulp cell, Inflammatory and immunological response, Myogenic differentiation Neuronal development, Odontoblast differentiation, Neuronal development, Odontoblast differentiation | ZEB2 | 5 |
|  |  | AFF3 |  |
|  |  | PRKACB |  |
|  |  | REEP1 |  |
|  |  | HS2ST1 |  |
| hsa-miR-200b-3p | Dental pulp repair and regeneration, Differentiation of dental pulp cell, Inflammatory and immunological response, Myogenic differentiation Neuronal development, Odontoblast differentiation, Neuronal development, Odontoblast differentiation | ZEB2 | 5 |
|  |  | AFF3 |  |
|  |  | PRKACB |  |
|  |  | REEP1 |  |
|  |  | HS2ST1 |  |
| hsa-miR-19b-3p | Inflammatory and response Tissue homeostasis | LRP2 | 5 |
|  |  | SEMA4C |  |
|  |  | EDARADD |  |
|  |  | PRKACB |  |
|  |  | WDR26 |  |
| hsa-let-7a-3p | Inflammatory and response Tissue homeostasis | CAMTA1 | 5 |
|  |  | DLX2 |  |
|  |  | CNNM4 |  |
|  |  | SPEN |  |
|  |  | COL11A1 |  |
| hsa-miR-218-5p | Dental pulp repair and regeneration, Differentiation of dental pulp cell, Inflammatory and immunological response, Myogenic differentiation Neuronal development, Odontoblast differentiation, Neuronal development, Odontoblast differentiation | KIF21B | 4 |
|  |  | CREB1 |  |
|  |  | ZEB2 |  |
|  |  | GALNT3 |  |
| hsa-miR-182-5p | Bone formation, Cell Survival Differentitation Inflammatory and immunological response Tooth movement | PRKACB | 4 |
|  |  | CAMTA1 |  |
|  |  | CREB1 |  |
|  |  | IGSF3 |  |
| hsa-miR-15a-5p | Inflammatory and response Tissue homeostasis | KIF1B | 4 |
|  |  | AKT3 |  |
|  |  | MFN2 |  |
|  |  | WNT3A |  |
|  |  | HSPG2 |  |
| hsa-miR-218-5p | Bone formation, Cell Survival Differentitation Inflammatory and immunological response Tooth movement | KIF21B | 4 |
|  |  | CREB1 |  |
|  |  | ZEB2 |  |
|  |  | GALNT3 |  |
| hsa-miR-424-5p | Dental pulp repair and regeneration, Differentiation of dental pulp cell, Inflammatory and immunological response, Myogenic differentiation Neuronal development, Odontoblast differentiation, Neuronal development, Odontoblast differentiation | KIF1B | 4 |
|  |  | AKT3 |  |
|  |  | MFN2 |  |
|  |  | HSPG2 |  |
| hsa-miR-141-3p | Dental pulp repair and regeneration, Differentiation of dental pulp cell, Inflammatory and immunological response, Myogenic differentiation Neuronal development, Odontoblast differentiation,Neuronal development,Odontoblast differentiation | ZEB2 | 3 |
|  |  | PRKACB |  |
|  |  | HS2ST1 |  |
| hsa-miR-200a-3p | Inflammatory and immunological response ,  Tooth movement | ZEB2 | 3 |
|  |  | PRKACB |  |
|  |  | HS2ST1 |  |
|  |  | ZEB2 |  |
|  |  | GALNT3 |  |
| hsa-miR-7-5p | Functional roles in tumor invasion and metastasis | SEMA4C | 3 |
|  |  | CNNM4 |  |
|  |  | GALNT3 |  |
| hsa-miR-23a-3p | Inflammatory and response Tissue homeostasis | SATB2 | 3 |
|  |  | CAMTA1 |  |
|  |  | REEP1 |  |
| hsa-miR-106b-5p | Inflammatory and response Tissue homeostasis | CAMTA1 | 3 |
|  |  | CTSK |  |
|  |  | CREB1 |  |
| hsa-miR-27a-3p | Involved in Epithelial Mesenchymal Transition and Mesenchymal Epithelial Transition EMT/MET | CREB1 | 3 |
|  |  | DYNC2LI1 |  |
|  |  | GALNT3 |  |
| hsa-miR-29a-3p | Involved in Epithelial Mesenchymal Transition and Mesenchymal Epithelial Transition EMT/MET | COL3A1 | 3 |
|  |  | COL11A1 |  |
|  |  | AKT3 |  |
| hsa-miR-221-3p | Involved in Epithelial Mesenchymal Transition and Mesenchymal Epithelial Transition EMT/MET | GNAI3 | 3 |
|  |  | GALNT3 |  |
|  |  | WDR35 |  |
| hsa-miR-34a-5p | Dental pulp repair and regeneration, Differentiation of dental pulp cell, Inflammatory and immunological response, Myogenic differentiation Neuronal development, Odontoblast differentiation,Neuronal development,Odontoblast differentiation | SATB2 | 2 |
|  |  | CAMTA1 |  |
| hsa-miR-146a-3p | Inflammatory and immunological response ,  Tooth movement | IGSF3 | 2 |
|  |  | FANCL |  |
| hsa-miR-29b-3p | Inflammatory and immunological response ,  Tooth movement | COL3A1 | 2 |
|  |  | COL11A1 |  |
|  |  | AKT3 |  |
| hsa-miR-214-3p | Inflammatory and response Tissue homeostasis | MFN2 | 2 |
|  |  | LRP2 |  |
| hsa-miR-30d-3p | Inflammatory and response Tissue homeostasis | ZEB2 | 2 |
|  |  | CREB1 |  |
| hsa-miR-30a-3p | Inflammatory and response Tissue homeostasis | ZEB2 | 2 |
|  |  | CREB1 |  |
| hsa-miR-520d-3p | Inflammatory and response Tissue homeostasis | HS2ST1 | 2 |
|  |  | LRP2 |  |
| hsa-miR-195-3p | Bone formation, Cell Survival Differentitation Inflammatory and immunological response Tooth movement | IRF6 | 2 |
|  |  | COL11A1 |  |
| hsa-miR-4328 | Bone formation, Cell Survival Differentitation Inflammatory and immunological response Tooth movement | GALNT3 | 2 |
|  |  | CKAP2L |  |
|  |  | CREB1 |  |
|  |  | MYSM1 |  |
| hsa-miR-16-1-3p | Functional roles in tumor invasion and metastasis | SPEN | 2 |
|  |  | TMEM237 |  |
| hsa-miR-126-5p | Functional roles in tumor invasion and metastasis | MDH1 | 2 |
|  |  | CSF1 |  |
| hsa-miR-214-3p | Functional roles in tumor invasion and metastasis | MFN2 | 2 |
|  |  | LRP2 |  |
| hsa-miR-520c-3p | Functional roles in tumor invasion and metastasis | HS2ST1 | 2 |
|  |  | LRP2 |  |
| hsa-miR-101-3p | Differentiation of dental follicle cells tooth eruption, | ARID1A | 1 |
| hsa-miR-155-3p | Dental pulp repair and regeneration, Differentiation of dental pulp cell, Inflammatory and immunological response, Myogenic differentiation Neuronal development, Odontoblast differentiation,Neuronal development,Odontoblast differentiation | GNAI3 | 1 |
| hsa-miR-516a-3p | Dental pulp repair and regeneration, Differentiation of dental pulp cell, Inflammatory and immunological response, Myogenic differentiation Neuronal development, Odontoblast differentiation,Neuronal development,Odontoblast differentiation | CSF1 | 1 |
| hsa-miR-766-5p | Dental pulp repair and regeneration, Differentiation of dental pulp cell, Inflammatory and immunological response, Myogenic differentiation Neuronal development, Odontoblast differentiation,Neuronal development,Odontoblast differentiation | PDE6D | 1 |
| hsa-miR-885-5p | Dental pulp repair and regeneration, Differentiation of dental pulp cell, Inflammatory and immunological response, Myogenic differentiation Neuronal development, Odontoblast differentiation,Neuronal development,Odontoblast differentiation | GALNT3 | 1 |
| hsa-miR-155-3p | Inflammatory and immunological response ,  Tooth movement | GNAI3 | 1 |
| hsa-miR-1260a | Inflammatory and immunological response ,  Tooth movement | SPEN | 1 |
| hsa-miR-1260b | Inflammatory and immunological response,  Tooth movement | SPEN | 1 |
| hsa-miR-203a-3p | Inflammatory and immunological response,  Tooth movement | HNRNPR | 1 |
| hsa-miR-103a-3p | Involved in Epithelial Mesenchymal Transition and Mesenchymal Epithelial Transition EMT/MET | IGSF3 | 1 |
| hsa-miR-130a-3p | Inflammatory and response Tissue homeostasis | ACVR1 | 1 |
| hsa-miR-22-3p | Inflammatory and response Tissue homeostasis | AKT3 | 1 |
| hsa-miR-301a-3p | Inflammatory and response Tissue homeostasis | ACVR1 | 1 |
| hsa-miR-142-3p | Inflammatory and response Tissue homeostasis | ZEB2 | 1 |
| hsa-miR-548a-3p | Inflammatory and response Tissue homeostasis | CAMTA1 | 1 |
| hsa-miR-138-2-3p | Bone formation, Cell Survival Differentitation Inflammatory and immunological response Tooth movement | MBD5 | 1 |
| hsa-miR-145-5p | Bone formation, Cell Survival Differentitation Inflammatory and immunological response Tooth movement | NRAS | 1 |
| hsa-miR-24-3p | Bone formation, Cell Survival Differentitation Inflammatory and immunological response Tooth movement | VANGL2 | 1 |
| hsa-miR-224-5p | Bone formation, Cell Survival Differentitation Inflammatory and immunological response Tooth movement | HOXD13 | 1 |
| hsa-miR-204-3p | Involved in Epithelial Mesenchymal Transition and Mesenchymal Epithelial Transition EMT/MET | VANGL2 | 1 |
| hsa-miR-10a-5p | Functional roles in tumor invasion and metastasis | CREB1 | 1 |
| hsa-miR-17-3p | Functional roles in tumor invasion and metastasis | AKT3 | 1 |
| hsa-miR-21-3p | Functional roles in tumor invasion and metastasis | HS2ST1 | 1 |
| hsa-miR-22-3p | Inflammatory and response Tissue homeostasis | AKT3 | 1 |
| hsa-miR-122b-5p | Functional roles in tumor invasion and metastasis | DVL1 | 1 |

**Table3: P-values of different genes with respect to their Control and patients.**

| **S.No.** | **Gene Name** | **P-Value** | **DEGs** |
| --- | --- | --- | --- |
| 1 | **miR-218-5p** | 0.0123 | Up regulation |
| 2 | **miR-15b-5p** | 0.0001 | Up regulation |
| 3 | **miR-200b-3p** | 0.0001 | Up regulation |
| 4 | **miR-let-7a-3p** | 0.0054 | Down regulation |
| 5 | **miR-19b-3p** | 0.1786 | Not major change |
| 6 | **miR-30d-3p** | 0.09 | Up regulation |
| 7 | **miR-141-3p** | 0.7297 | Not major change |
| 8 | **miR-335-3p** | 0.805 | Not major change |

**Table4: miRNA Primer Design for Stem-Loop RT-qPCR Analysis.**

| **Gene primer name** | **5'<-----Sequence----->3'** |
| --- | --- |
| **miR-218-5P_SL** | GTCGTATCCAGTGCAGGGTCCGAGGTATTCGCACTGGATACGACACATGG |
| **miR-218-5P_FP** | AACACGCTTGTGCTTGATCT |
| **miR-15b-5p_SL** | GTCGTATCCAGTGCAGGGTCCGAGGTATTCGCACTGGATACGACTGTAAA |
| **miR-15b-5p_FP** | AACCGGTAGCAGCACATCAT |
| **miR-30d-3p_SL** | GTCGTATCCAGTGCAGGGTCCGAGGTATTCGCACTGGATACGACGCAGCA |
| **miR-30d-3p_FP** | AACACGCCTTTCAGTCAGATG |
| **miR-141-3P_SL** | GTCGTATCCAGTGCAGGGTCCGAGGTATTCGCACTGGATACGACCCATCT |
| **miR-141-3P_FP** | AACACGCTAACACTGTCTGGT |
| **miR-19b-3P_SL** | GTCGTATCCAGTGCAGGGTCCGAGGTATTCGCACTGGATACGACTCAGTT |
| **miR-19b-3P_FP** | AACAAGTGTGCAAATCCATGC |
| **miR-335-3p_SL** | GTCGTATCCAGTGCAGGGTCCGAGGTATTCGCACTGGATACGACGGTCAG |
| **miR-335-3p_FP** | AGCCAGCGTTTTTCATTATTGC |
| **miR-let-7a-3p_SL** | GTCGTATCCAGTGCAGGGTCCGAGGTATTCGCACTGGATACGACGAAAGA |
| **miR-let-7a-3p_FP** | ATGCGCGCCTATACAATCTAC |
| **miR-200b-3p_SL** | GTCGTATCCAGTGCAGGGTCCGAGGTATTCGCACTGGATACGACTCATCA |
| **MiR-200B-3P_FP** | AACCGGTAATACTGCCTGGT |
| **RNU6 FP** | CTCGCTTCGGCAGCACA |
| **RNU6 RP** | AACGCTTCACGAATTTGCGT |

**Table5:** Table and listing the metabolites from different resources

| **Metabolites associated with Diseases** | **Keywords search in DB** | **List of Metabolites** | | **Bio specimen** | **References** |
| --- | --- | --- | --- | --- | --- |
| Pulp repair by regulating inflammation and stimulating dentin regeneration in dental pulp stem cells |  | omega-3 polyunsaturated fatty-acid | | Dental pulp stem cell | Chen et al.2021 |
| Tooth Diseases | Tooth (under Diseases) | 2-Hydroxy-3+D14:D25-methylpentanoic acid | | Saliva | HMDB |
| Tooth Diseases | Tooth (uder Diseases) | 4-Hydroxybutyric acid | | Saliva | HMDB |
| Tooth Diseases | Tooth (under Diseases) | 4-Hydroxyproline | | Saliva | HMDB |
| Tooth Diseases | Tooth (under Diseases) | Citrulline | | Saliva | HMDB |
| Tooth Diseases | Tooth (under Diseases) | Cortisone | | Saliva | HMDB |
| Tooth Diseases | Tooth (under Diseases) | Guanosine | | Saliva | HMDB |
| Tooth Diseases | Tooth (under Diseases) | Hydrocinnamic acid | | Saliva | HMDB |
| Tooth Diseases | Tooth (under Diseases) | Hydroxyisocaproic acid | | Saliva | HMDB |
| Tooth Diseases | Tooth (under Diseases) | Indoleacetic acid | | Saliva | HMDB |
| Tooth Diseases | Tooth (under Diseases) | Inosine | | Saliva | HMDB |
| Tooth Diseases | Tooth (under Diseases) | Isovaleric acid | | Saliva | HMDB |
| Tooth Diseases | Tooth (under Diseases) | Ketoleucine | | Saliva | HMDB |
| Tooth Diseases | Tooth (Metabolites) | [Doxycycline](https://hmdb.ca/metabolites/HMDB0014399) | | Urine | HMDB |
| Tooth Diseases | Tooth (Metabolites) |  | | Urine | HMDB |
|  |  | [Hernandulcin](https://hmdb.ca/metabolites/HMDB0037906) |  |  |  |
| Tooth Diseases | Tooth (Metabolites) | [Maltitol](https://hmdb.ca/metabolites/HMDB0002928) | | Urine | HMDB |
| Tooth Diseases | Tooth (Metabolites) | [Deoxythymidine diphosphate-L-rhamnose](https://hmdb.ca/metabolites/HMDB0006354) | | Urine | HMDB |
| Tooth Diseases | Tooth (Metabolites) |  | | Urine | HMDB |
|  |  | [Erythritol](https://hmdb.ca/metabolites/HMDB0002994) |  |  |  |
| Tooth Diseases | Tooth (Metabolites) |  | | Urine | HMDB |
|  |  | [Calcium](https://hmdb.ca/metabolites/HMDB0000464) | |  |  |
| Tooth Diseases | Tooth (Metabolites) | [Hydrogen peroxide](https://hmdb.ca/metabolites/HMDB0003125) | | Urine | HMDB |
| Tooth Diseases | Tooth (Metabolites) | [Doxycycline](https://hmdb.ca/metabolites/HMDB0014399) | | Urine | HMDB |
| Tooth Diseases | Tooth (Metabolites) |  | | Urine | HMDB |
|  |  | [Hernandulcin](https://hmdb.ca/metabolites/HMDB0037906) | |  |  |
| Tooth Diseases | Tooth (Metabolites) | [Maltitol](https://hmdb.ca/metabolites/HMDB0002928) | | Urine | HMDB |
| Tooth Diseases | Tooth Enamel (Metabolite) | [Erythritol](https://hmdb.ca/metabolites/HMDB0002994) | | Blood, Urine, Saliva | HMDB |
| Tooth Diseases | Tooth Enamel (Metabolite) | Silux | | Blood | HMDB |
| Tooth Diseases | Tooth Enamel (Diseases) | 2-Hydroxy-3-methylpentanoic | | Saliva | HMDB |
| Tooth Diseases | Tooth Enamel (Diseases) | 4-Hydroxybutyric | | Saliva | HMDB |
| Tooth Diseases | Tooth Enamel (Diseases) | 4-Hydroxyproline | | Saliva | HMDB |
| Tooth Diseases | Tooth Enamel (Diseases) | Citrulline | | Saliva | HMDB |
| Tooth Diseases | Tooth Enamel (Diseases) | Cortisone | | Saliva | HMDB |
| Tooth Diseases | Tooth Enamel (Diseases) | Guanosine | | Saliva | HMDB |
| Tooth Diseases | Tooth Enamel (Diseases) | Hydrocinnamic | | Saliva | HMDB |
| Tooth Diseases | Tooth Enamel (Diseases) | Hydroxyisocaproic | | Saliva | HMDB |
| Tooth Diseases | Tooth Enamel (Diseases) | Indoleacetic | | Saliva | HMDB |
| Tooth Diseases | Tooth Enamel (Diseases) | Inosine | | Saliva | HMDB |
| Tooth Diseases | Tooth Enamel (Diseases) | Isovaleric | | Saliva | HMDB |
| Tooth Diseases | Tooth Enamel (Diseases) | Ketoleucine | | Saliva | HMDB |
| Tooth Diseases | Tooth Enamel (Diseases) | L-Acetylcarnitine | | Saliva | HMDB |
| Tooth Diseases | Tooth Enamel (Diseases) | L-Carnitine | | Saliva | HMDB |
| Tooth Diseases | Tooth Enamel (Diseases) | L-Proline | | Saliva | HMDB |
| Tooth Diseases | Tooth Enamel (Diseases) | Mevalonic | | Saliva | HMDB |
| Tooth Diseases | Tooth Enamel (Diseases) | Phenylacetic | | Saliva | HMDB |
| Tooth Diseases | Tooth Enamel (Diseases) | Propionylcarnitine | | Saliva | HMDB |
| Tooth Diseases | Tooth Enamel (Diseases) | Spermidine | | Saliva | HMDB |
| Bone Diseses |  | [Methyl methacrylate](https://hmdb.ca/metabolites/HMDB0032385) | | Blood, Saliva, Urine | HMDB |
|  | Bone (Metabolite) |  |  |  |  |
| Bone Diseses |  |  | | Blood, Saliva, Urine | HMDB |
|  | Bone (Metabolite) | [Ibandronate](https://hmdb.ca/metabolites/HMDB0014848) |  |  |  |
| Bone Diseses |  | [Etidronic acid](https://hmdb.ca/metabolites/HMDB0015210) | | Blood, Saliva, Urine | HMDB |
|  | Bone (Metabolite) |  |  |  |  |
| Bone Diseses |  | [Galactosylhydroxylysine](https://hmdb.ca/metabolites/HMDB0000600) | | Blood, Saliva, Urine | HMDB |
|  | Bone (Metabolite) |  |  |  |  |
| Bone Diseses |  | [Pamidronate](https://hmdb.ca/metabolites/HMDB0014427) | | Blood, Saliva, Urine | HMDB |
|  | Bone (Metabolite) |  |  |  |  |
| Bone Diseses |  | [Deoxypyridinoline](https://hmdb.ca/metabolites/HMDB0000569) | | Blood, Saliva, Urine | HMDB |
|  | Bone (Metabolite) |  |  |  |  |
| Bone Diseses |  | [Tiludronate](https://hmdb.ca/metabolites/HMDB0015265) | | Blood, Saliva, Urine | HMDB |
|  | Bone (Metabolite) |  |  |  |  |
| Bone Diseses |  | [Risedronate](https://hmdb.ca/metabolites/HMDB0015022) | | Blood, Saliva, Urine | HMDB |
|  | Bone (Metabolite) |  |  |  |  |
| Bone Diseses |  | [Raloxifene](https://hmdb.ca/metabolites/HMDB0014624) | | Blood, Saliva, Urine | HMDB |
|  | Bone (Metabolite) |  |  |  |  |
| Bone Diseses |  | [Clodronate](https://hmdb.ca/metabolites/HMDB0014858) | | Blood, Saliva, Urine | HMDB |
|  | Bone (Metabolite) |  |  |  |  |
| Bone Diseses |  | [Pyridinoline](https://hmdb.ca/metabolites/HMDB0000851) | | Blood, Saliva, Urine | HMDB |
|  | Bone (Metabolite) |  |  |  |  |
| Bone Diseses |  | [24-Hydroxycalcitriol](https://hmdb.ca/metabolites/HMDB0006228) | | Blood, Saliva, Urine | HMDB |
|  | Bone (Metabolite) |  |  |  |  |
| Bone Diseses |  | [Silicon](https://hmdb.ca/metabolites/HMDB0002175) | | Blood, Saliva, Urine | HMDB |
|  | Bone (Metabolite) |  |  |  |  |
| Bone Diseses |  | [19-Hydroxy-PGE2](https://hmdb.ca/metabolites/HMDB0001908) | | Blood, Saliva, Urine | HMDB |
|  | Bone (Metabolite) |  |  |  |  |
| Bone Diseses |  | [15-Keto-13,14-dihydroprostaglandin A2](https://hmdb.ca/metabolites/HMDB0001244) | | Blood, Saliva, Urine | HMDB |
|  | Bone (Metabolite) |  |  |  |  |
| Bone Diseses |  | [Alfacalcidol](https://hmdb.ca/metabolites/HMDB0015504) | | Blood, Saliva, Urine | HMDB |
|  | Bone (Metabolite) |  |  |  |  |
| Bone Diseses |  | [Calcitroic acid](https://hmdb.ca/metabolites/HMDB0006472) | | Blood, Saliva, Urine | HMDB |
|  | Bone (Metabolite) |  |  |  |  |
| Bone Diseses |  | [Zoledronate](https://hmdb.ca/metabolites/HMDB0014543) | | Blood, Saliva, Urine | HMDB |
|  | Bone (Metabolite) |  |  |  |  |
| Bone Diseses |  | [Prolylhydroxyproline](https://hmdb.ca/metabolites/HMDB0006695) | | Blood, Saliva, Urine | HMDB |
|  | Bone (Metabolite) |  |  |  |  |
| Bone Diseses |  | [Indoxyl sulfate](https://hmdb.ca/metabolites/HMDB0000682) | | Blood, Saliva, Urine | HMDB |
|  | Bone (Metabolite) |  |  |  |  |
| Bone Diseses |  | [15-Keto-prostaglandin E2](https://hmdb.ca/metabolites/HMDB0003175) | | Blood, Saliva, Urine | HMDB |
|  | Bone (Metabolite) |  |  |  |  |
| Bone Diseses |  | [Prostaglandin E2](https://hmdb.ca/metabolites/HMDB0001220) | | Blood, Saliva, Urine | HMDB |
|  | Bone (Metabolite) |  |  |  |  |
| Bone Diseses |  | [Pyrophosphate](https://hmdb.ca/metabolites/HMDB0000250) | | Blood, Saliva, Urine | HMDB |
|  | Bone (Metabolite) |  |  |  |  |
| Bone Diseses |  | [Strontium](https://hmdb.ca/metabolites/HMDB0003642) | | Blood, Saliva, Urine | HMDB |
|  | Bone (Metabolite) |  |  |  |  |
| Bone Diseses |  | [Tridecanol](https://hmdb.ca/metabolites/HMDB0013316) | | Blood, Saliva, Urine | HMDB |
|  | Bone (Metabolite) |  |  |  |  |
| Bone Diseses |  | [Vitamin A](https://hmdb.ca/metabolites/HMDB0000305) | | Blood, Urine | HMDB |
|  | Bone (Metabolite) |  |  |  |  |
| Bone Diseses |  |  | | Blood, Urine | HMDB |
|  | Bone (Metabolite) | [20-Hydroxy-PGE2](https://hmdb.ca/metabolites/HMDB0003247) |  |  |  |
| Bone Diseses |  | [Demethylphylloquinone](https://hmdb.ca/metabolites/HMDB0004649) | | Blood, Urine | HMDB |
|  | Bone (Metabolite) |  |  |  |  |
| Bone Diseses |  | [Alendronic acid](https://hmdb.ca/metabolites/HMDB0001915) | | Blood, Urine | HMDB |
|  | Bone (Metabolite) |  |  |  |  |
| Bone Diseses |  | [Melphalan](https://hmdb.ca/metabolites/HMDB0015176) | | Blood, Urine | HMDB |
|  | Bone (Metabolite) |  |  |  |  |
| Bone Diseses |  | [13,14-Dihydro-15-keto-PGE2](https://hmdb.ca/metabolites/HMDB0002776) | | Blood, Urine | HMDB |
|  | Bone (Metabolite) |  |  |  |  |
| Bone Diseses |  | [Spermine dialdehyde](https://hmdb.ca/metabolites/HMDB0013076) | | Blood, Urine | HMDB |
|  | Bone (Metabolite) |  |  |  |  |
| Bone Diseses |  |  | | Blood, Urine | HMDB |
|  | Bone (Metabolite) | [8-isoprostaglandin E2](https://hmdb.ca/metabolites/HMDB0005844) |  |  |  |
| Bone Diseses |  |  | | Blood, Urine | HMDB |
|  | Bone (Metabolite) | [8-iso-15-keto-PGE2](https://hmdb.ca/metabolites/HMDB0002341) |  |  |  |
| Bone Diseses |  | [Calcitriol](https://hmdb.ca/metabolites/HMDB0001903) | | Blood, Urine | HMDB |
|  | Bone (Metabolite) |  |  |  |  |
| Bone Diseses |  | [Manganese](https://hmdb.ca/metabolites/HMDB0001333) | | Blood, Urine | HMDB |
|  | Bone (Metabolite) |  |  |  |  |
| Bone Diseses |  | [2,6-Dimethylpyridine](https://hmdb.ca/metabolites/HMDB0032972) | | Blood, Urine | HMDB |
|  | Bone (Metabolite) |  |  |  |  |
| Bone Diseses |  | [Thiotepa](https://hmdb.ca/metabolites/HMDB0015576) | | Blood, Urine | HMDB |
|  | Bone (Metabolite) |  |  |  |  |
| Bone Diseses |  | [Busulfan](https://hmdb.ca/metabolites/HMDB0015143) | | Blood, Urine | HMDB |
|  | Bone (Metabolite) |  |  |  |  |
| Bone Diseses |  | [5-Hydroxylysine](https://hmdb.ca/metabolites/HMDB0000450) | | Blood, Urine | HMDB |
|  | Bone (Metabolite) |  |  |  |  |
| Bone Diseses |  | [Uracil mustard](https://hmdb.ca/metabolites/HMDB0014929) | | Blood, Urine | HMDB |
|  | Bone (Metabolite) |  |  |  |  |
| Bone Diseses |  | [13-Demethyl tacrolimus](https://hmdb.ca/metabolites/HMDB0060706) | | Blood, Urine | HMDB |
|  | Bone (Metabolite) |  |  |  |  |
| Bone Diseses |  | [Bafetinib](https://hmdb.ca/metabolites/HMDB0240206) | | Blood, Urine | HMDB |
|  | Bone (Metabolite) |  |  |  |  |
| Bone Diseses |  | [Mitotane](https://hmdb.ca/metabolites/HMDB0014786) | | Blood, Urine | HMDB |
|  | Bone (Metabolite) |  |  |  |  |
| Bone Diseses |  | [31-O-Demethyltacrolimus](https://hmdb.ca/metabolites/HMDB0061049) | | Blood, Urine | HMDB |
|  | Bone (Metabolite) |  |  |  |  |
| Bone Diseses |  | [Tin](https://hmdb.ca/metabolites/HMDB0001960) | | Blood, Urine | HMDB |
|  | Bone (Metabolite) |  |  |  |  |
| Bone Diseses |  | [1,25-Dihydroxyvitamin D3-26,23-lactone](https://hmdb.ca/metabolites/HMDB0000969) | | Blood, Urine | HMDB |
|  | Bone (Metabolite) |  |  |  |  |
| Bone Diseses |  | [Lindane](https://hmdb.ca/metabolites/HMDB0014575) | | Blood, Urine | HMDB |
|  | Bone (Metabolite) |  |  |  |  |
| Bone Diseses |  | [Plicamycin](https://hmdb.ca/metabolites/HMDB0015682) | | Blood, Urine | HMDB |
|  | Bone (Metabolite) |  |  |  |  |
| Bone Diseses |  | [Nivalenol](https://hmdb.ca/metabolites/HMDB0004304) | | Blood, Urine | HMDB |
|  | Bone (Metabolite) |  |  |  |  |
| Bone Diseses |  | [Zidovudine](https://hmdb.ca/metabolites/HMDB0014638) | | Blood, Urine, Saliva | HMDB |
|  | Bone (Metabolite) |  |  |  |  |
| Bone Diseses |  |  | | Blood, Urine, Saliva | HMDB |
|  | Bone (Metabolite) | [Cefonicid](https://hmdb.ca/metabolites/HMDB0015423) |  |  |  |
| Bone Diseses |  | [6-beta-hydrocortisol](https://hmdb.ca/metabolites/HMDB0061033) | | Blood, Urine, Saliva | HMDB |
|  | Bone (Metabolite) |  |  |  |  |
| Bone Diseses |  | [Plerixafor](https://hmdb.ca/metabolites/HMDB0015681) | | Blood, Urine, Saliva | HMDB |
|  | Bone (Metabolite) |  |  |  |  |
| Bone Diseses |  | [Calcium](https://hmdb.ca/metabolites/HMDB0000464) | | Blood, Urine, Saliva | HMDB |
|  | Bone (Metabolite) |  |  |  |  |
| Bone Diseses |  | [Resolvin E1](https://hmdb.ca/metabolites/HMDB0010410) | | Blood, Urine, Saliva | HMDB |
|  | Bone (Metabolite) |  |  |  |  |
| Bone Diseses |  | [Testosterone enanthate](https://hmdb.ca/metabolites/HMDB0005814) | | Blood, Urine, Saliva | HMDB |
|  | Bone (Metabolite) |  |  |  |  |
| Bone Diseses |  | [N-Ethylglycine](https://hmdb.ca/metabolites/HMDB0041945) | | Blood, Urine, Saliva | HMDB |
|  | Bone (Metabolite) |  |  |  |  |
| Bone Diseses |  | [Auranofin](https://hmdb.ca/metabolites/HMDB0015130) | | Blood, Urine, Saliva | HMDB |
|  | Bone (Metabolite) |  |  |  |  |
| Bone Diseses |  | [Tacrolimus](https://hmdb.ca/metabolites/HMDB0015002) | | Blood, Urine, Saliva | HMDB |
|  | Bone (Metabolite) |  |  |  |  |
| Bone Diseses |  | [Dihydrotachysterol](https://hmdb.ca/metabolites/HMDB0015203) | | Blood, Urine, Saliva | HMDB |
|  | Bone (Metabolite) |  |  |  |  |
| Bone Diseses |  | N-Acetyl-D-glucosamine | | Blood, Urine, Saliva | HMDB |
|  | Bone (Metabolite) |  |  |  |  |
| Bone Diseses |  | [Calcipotriol](https://hmdb.ca/metabolites/HMDB0015567) | | Blood, Urine, Saliva | HMDB |
|  | Bone (Metabolite) |  |  |  |  |
| Bone Diseses |  | [Mechlorethamine](https://hmdb.ca/metabolites/HMDB0015025) | | Blood, Urine, Saliva | HMDB |
|  | Bone (Metabolite) |  |  |  |  |
| Bone Diseses |  | [Phosphate](https://hmdb.ca/metabolites/HMDB0001429) | | Blood, Urine, Saliva | HMDB |
|  | Bone (Metabolite) |  |  |  |  |
| Bone Diseses |  | [Tamoxifen](https://hmdb.ca/metabolites/HMDB0014813) | | Blood, Urine, Saliva | HMDB |
|  | Bone (Metabolite) |  |  |  |  |
| Bone Diseses |  | [23S,25-dihydroxyvitamin D3](https://hmdb.ca/metabolites/HMDB0006720) | | Blood, Urine, Saliva | HMDB |
|  | Bone (Metabolite) |  |  |  |  |
| Bone Diseses |  | [Guggulsterone](https://hmdb.ca/metabolites/HMDB0002726) | | Blood, Urine, Saliva | HMDB |
|  | Bone (Metabolite) |  |  |  |  |
| Bone Diseses |  | [Toremifene](https://hmdb.ca/metabolites/HMDB0014679) | | Blood, Urine, Saliva | HMDB |
|  | Bone (Metabolite) |  |  |  |  |
| Bone Diseses |  | [Azacitidine](https://hmdb.ca/metabolites/HMDB0015063) | | Blood, Urine, Saliva | HMDB |
|  | Bone (Metabolite) |  |  |  |  |
| Bone Diseses |  | [Beta-Aminopropionitrile](https://hmdb.ca/metabolites/HMDB0004101) | | Blood, Urine, Saliva | HMDB |
|  | Bone (Metabolite) |  |  |  |  |
| Bone Diseses |  | [24,25-Dihydroxyvitamin D](https://hmdb.ca/metabolites/HMDB0000430) | | Blood, Urine, Saliva | HMDB |
|  | Bone (Metabolite) |  |  |  |  |
| Bone Diseses |  | [SM(d18:1/12:0)](https://hmdb.ca/metabolites/HMDB0012096) | | Blood, Urine, Saliva | HMDB |
|  | Bone (Metabolite) |  |  |  |  |
| Bone Diseses |  | [Homo-L-arginine](https://hmdb.ca/metabolites/HMDB0000670) | | Blood, Urine, Saliva | HMDB |
|  | Bone (Metabolite) |  |  |  |  |
| Bone Diseses |  | [Hydroxysphingomyeline C22:1](https://hmdb.ca/metabolites/HMDB0013466) | | Blood, Urine, Saliva | HMDB |
|  | Bone (Metabolite) |  |  |  |  |
| Bone Diseses |  | [SM(d16:1/20:3(8Z,11Z,14Z)-O(5,6))](https://hmdb.ca/metabolites/HMDB0290254) | | NA | HMDB |
|  | Bone (Metabolite) |  |  |  |  |
| Bone Diseses |  | [SM(d16:1/6 keto-PGF1alpha)](https://hmdb.ca/metabolites/HMDB0290267) |  | NA | HMDB |
|  | Bone (Metabolite) |  |  |  |  |
| Bone Diseses |  | [SM(d16:1/PGF2alpha)](https://hmdb.ca/metabolites/HMDB0290268) | | NA | HMDB |
|  | Bone (Metabolite) |  |  |  |  |
| Bone Diseses |  | [SM(d16:1/5-iso PGF2VI)](https://hmdb.ca/metabolites/HMDB0290274) | | NA | HMDB |
|  | Bone (Metabolite) |  |  |  |  |
| Bone Diseses |  | [SM(d16:1/18:2(10E,12Z)+=O(9))](https://hmdb.ca/metabolites/HMDB0290277) | | NA | HMDB |
|  | Bone (Metabolite) |  |  |  |  |
| Bone Diseses |  | [SM(d16:1/18:1(12Z)-2OH(9,10))](https://hmdb.ca/metabolites/HMDB0290279) | | NA | HMDB |
|  | Bone (Metabolite) |  |  |  |  |
| Bone Diseses |  | [SM(d16:1/18:1(12Z)-O(9S,10R))](https://hmdb.ca/metabolites/HMDB0290280) | | NA | HMDB |
|  | Bone (Metabolite) |  |  |  |  |
| Bone Diseses |  | [SM(d16:1/18:3(9,11,15)-OH(13))](https://hmdb.ca/metabolites/HMDB0290289) | | NA | HMDB |
|  | Bone (Metabolite) |  |  |  |  |
| Bone Diseses |  | [SM(d16:2(4E,8Z)/18:1(9Z)-O(12,13))](https://hmdb.ca/metabolites/HMDB0290325) | | NA | HMDB |
|  | Bone (Metabolite) |  |  |  |  |
| Bone Diseses |  | [SM(d16:2(4E,8Z)/18:3(9,11,15)-OH(13))](https://hmdb.ca/metabolites/HMDB0290341) | | NA | HMDB |
|  | Bone (Metabolite) |  |  |  |  |
| Bone Diseses |  | [SM(d16:2(4E,8Z)/20:3(6,8,11)-OH(5))](https://hmdb.ca/metabolites/HMDB0290345) | | NA | HMDB |
|  | Bone (Metabolite) |  |  |  |  |
| Bone Diseses |  | [SM(d17:1/20:3(8Z,11Z,14Z)-2OH(5,6))](https://hmdb.ca/metabolites/HMDB0290369) | | NA | HMDB |
|  | Bone (Metabolite) |  |  |  |  |
| Bone Diseses |  | [SM(d17:1/5-iso PGF2VI)](https://hmdb.ca/metabolites/HMDB0290370) | | NA | HMDB |
|  | Bone (Metabolite) |  |  |  |  |
| Bone Diseses |  | [SM(d17:1/18:3(10,12,15)-OH(9))](https://hmdb.ca/metabolites/HMDB0290392) | | NA | HMDB |
|  | Bone (Metabolite) |  |  |  |  |
| Bone Diseses |  | [SM(d17:1/PGE1)](https://hmdb.ca/metabolites/HMDB0290394) | | NA | HMDB |
|  | Bone (Metabolite) |  |  |  |  |
| Bone Diseses |  | [SM(d17:2(4E,8Z)/PGJ2)](https://hmdb.ca/metabolites/HMDB0290416) | | NA | HMDB |
|  | Bone (Metabolite) |  |  |  |  |
| Bone Diseses |  | [SM(d17:2(4E,8Z)/18:1(9Z)-O(12,13))](https://hmdb.ca/metabolites/HMDB0290429) | | NA | HMDB |
|  | Bone (Metabolite) |  |  |  |  |
| Bone Diseses |  | [SM(d18:0/PGJ2)](https://hmdb.ca/metabolites/HMDB0290468) | | NA | HMDB |
|  | Bone (Metabolite) |  |  |  |  |
| Bone Diseses |  | [SM(d18:0/20:3(8Z,11Z,14Z)-2OH(5,6))](https://hmdb.ca/metabolites/HMDB0290473) | | NA | HMDB |
|  | Bone (Metabolite) |  |  |  |  |
| Bone Diseses |  | [SM(d18:0/18:1(12Z)-2OH(9,10))](https://hmdb.ca/metabolites/HMDB0290479) | | NA | HMDB |
|  | Bone (Metabolite) |  |  |  |  |
| Bone Diseses |  | [SM(d18:0/PGD1)](https://hmdb.ca/metabolites/HMDB0290499) | | NA | HMDB |
|  | Bone (Metabolite) |  |  |  |  |
| Bone Diseses |  | [SM(d18:1/PGE2)](https://hmdb.ca/metabolites/HMDB0290516) | | NA | HMDB |
|  | Bone (Metabolite) |  |  |  |  |
| Bone Diseses | Bone (Metabolite) | [SM(d19:0/5-iso PGF2VI)](https://hmdb.ca/metabolites/HMDB0290630) | | NA | HMDB |
| Bone Diseses | Bone (Metabolite) | [SM(d19:0/18:1(9Z)-O(12,13))](https://hmdb.ca/metabolites/HMDB0290637) |  | NA | HMDB |
| Bone Diseses | Bone (Metabolite) | [SM(d19:0/18:3(10,12,15)-OH(9))](https://hmdb.ca/metabolites/HMDB0290652) | | NA | HMDB |
| Bone Diseses | Bone (Metabolite) | [SM(d19:1/PGF2alpha)](https://hmdb.ca/metabolites/HMDB0290675) | | NA | HMDB |
| Bone Diseses | Bone (Metabolite) | [SM(d19:1/TXB2)](https://hmdb.ca/metabolites/HMDB0290679) | | NA | HMDB |
| Bone Diseses | Bone (Metabolite) | [SM(d19:1/18:2(9Z,11E)+=O(13))](https://hmdb.ca/metabolites/HMDB0290686) | | NA | HMDB |
| Bone Diseses | Bone (Metabolite) | [SM(d19:1/18:3(9,11,15)-OH(13))](https://hmdb.ca/metabolites/HMDB0290705) | | NA | HMDB |
| Bone Diseses | Bone (Metabolite) | [SM(d20:1/LTE4)](https://hmdb.ca/metabolites/HMDB0290729) | | NA | HMDB |
| Bone Diseses | Bone (Metabolite) | [SM(d20:1/TXB2)](https://hmdb.ca/metabolites/HMDB0290731) | | NA | HMDB |
| Bone Diseses | Bone (Metabolite) | [SM(d20:1/20:3(8Z,11Z,14Z)-2OH(5,6))](https://hmdb.ca/metabolites/HMDB0290733) | | NA | HMDB |
| Bone Diseses | Bone (Metabolite) | [SM(d20:1/18:2(10E,12Z)+=O(9))](https://hmdb.ca/metabolites/HMDB0290737) | | NA | HMDB |
| Bone Diseses | Bone (Metabolite) | [Vanadium](https://hmdb.ca/metabolites/HMDB0002503) | | NA | HMDB |
| Bone Diseses | Bone (Metabolite) | [SM(d18:0/18:1(11Z))](https://hmdb.ca/metabolites/HMDB0012088) | | NA | HMDB |
| Bone Diseses | Bone (Metabolite) | [SM(d18:0/22:1(13Z))](https://hmdb.ca/metabolites/HMDB0012092) | | NA | HMDB |
| Bone Diseses | Bone (Metabolite) | [SM(d18:1/18:1)](https://hmdb.ca/metabolites/HMDB0012100) | | NA | HMDB |
| Bone Diseses | Bone (Metabolite) | [Hydroxysphingomyeline C24:1](https://hmdb.ca/metabolites/HMDB0013469) | | NA | HMDB |
| Bone Diseses | Bone (Metabolite) | [SM(d16:1/20:3(5Z,11Z,14Z)-O(8,9))](https://hmdb.ca/metabolites/HMDB0290253) | | NA | HMDB |
| Bone Diseses | Bone (Metabolite) | [SM(d16:1/TXB2)](https://hmdb.ca/metabolites/HMDB0290271) | | NA | HMDB |
| Bone Diseses | Bone (Metabolite) | [SM(d16:1/18:2(9Z,11E)+=O(13))](https://hmdb.ca/metabolites/HMDB0290278) | | NA | HMDB |
| Bone Diseses | Bone (Metabolite) | [SM(d16:2(4E,8Z)/PGF2alpha)](https://hmdb.ca/metabolites/HMDB0290311) | | NA | HMDB |
| Bone Diseses | Bone (Metabolite) | [SM(d16:2(4E,8Z)/5-iso PGF2VI)](https://hmdb.ca/metabolites/HMDB0290318) | | NA | HMDB |
| Bone Diseses | Bone (Metabolite) | [SM(d17:1/20:3(8Z,11Z,14Z)-O(5,6))](https://hmdb.ca/metabolites/HMDB0290349) | | NA | HMDB |
| Bone Diseses | Bone (Metabolite) | [SM(d17:1/LTE4)](https://hmdb.ca/metabolites/HMDB0290365) | | NA | HMDB |
| Bone Diseses | Bone (Metabolite) | [SM(d17:1/18:1(9Z)-O(12,13))](https://hmdb.ca/metabolites/HMDB0290377) | | NA | HMDB |
| Bone Diseses | Bone (Metabolite) | [SM(d17:2(4E,8Z)/PGD2)](https://hmdb.ca/metabolites/HMDB0290413) | | NA | HMDB |
| Bone Diseses | Bone (Metabolite) | [SM(d17:2(4E,8Z)/PGF2alpha)](https://hmdb.ca/metabolites/HMDB0290415) | | NA | HMDB |
| Bone Diseses | Bone (Metabolite) | [SM(d18:0/20:3(8Z,11Z,14Z)-O(5,6))](https://hmdb.ca/metabolites/HMDB0290453) |  | NA | HMDB |
| Bone Diseses | Bone (Metabolite) | [SM(d18:0/PGD2)](https://hmdb.ca/metabolites/HMDB0290465) | | NA | HMDB |
| Bone Diseses | Bone (Metabolite) | [SM(d18:0/LTE4)](https://hmdb.ca/metabolites/HMDB0290469) | | NA | HMDB |
| Bone Diseses | Bone (Metabolite) | [SM(d18:0/18:3(10,12,15)-OH(9))](https://hmdb.ca/metabolites/HMDB0290496) | | NA | HMDB |
| Bone Diseses | Bone (Metabolite) | [SM(d18:0/PGF1alpha)](https://hmdb.ca/metabolites/HMDB0290500) | | NA | HMDB |
| Bone Diseses | Bone (Metabolite) | [SM(d18:1/PGD1)](https://hmdb.ca/metabolites/HMDB0290551) | | NA | HMDB |
| Bone Diseses | Bone (Metabolite) | [SM(d18:2(4E,14Z)/18:1(9Z)-O(12,13))](https://hmdb.ca/metabolites/HMDB0290585) | | NA | HMDB |
| Bone Diseses | Bone (Metabolite) | [SM(d18:2(4E,14Z)/18:3(10,12,15)-OH(9))](https://hmdb.ca/metabolites/HMDB0290600) | | NA | HMDB |
| Bone Diseses | Bone (Metabolite) | [SM(d18:2(4E,14Z)/18:3(9,11,15)-OH(13))](https://hmdb.ca/metabolites/HMDB0290601) | | NA | HMDB |
| Bone Diseses | Bone (Metabolite) | [SM(d19:0/18:2(10E,12Z)+=O(9))](https://hmdb.ca/metabolites/HMDB0290633) | | NA | HMDB |
| Bone Diseses | Bone (Metabolite) | [SM(d19:0/18:1(12Z)-O(9S,10R))](https://hmdb.ca/metabolites/HMDB0290636) | | NA | HMDB |
| Bone Diseses | Bone (Metabolite) | [SM(d19:0/PGE1)](https://hmdb.ca/metabolites/HMDB0290654) | | NA | HMDB |
| Bone Diseses | Bone (Metabolite) | [SM(d19:1/20:3(5Z,11Z,14Z)-O(8,9))](https://hmdb.ca/metabolites/HMDB0290660) | | NA | HMDB |
| Bone Diseses | Bone (Metabolite) | [SM(d19:1/PGE2)](https://hmdb.ca/metabolites/HMDB0290672) | | NA | HMDB |
| Bone Diseses | Bone (Metabolite) | [SM(d19:1/18:3(10,12,15)-OH(9))](https://hmdb.ca/metabolites/HMDB0290704) | | NA | HMDB |
| Bone Diseses | Bone (Metabolite) | [SM(d19:1/20:3(6,8,11)-OH(5))](https://hmdb.ca/metabolites/HMDB0290709) | | NA | HMDB |
| Bone Diseses | Bone (Metabolite) | [SM(d20:1/20:3(8Z,11Z,14Z)-O(5,6))](https://hmdb.ca/metabolites/HMDB0290713) | | NA | HMDB |
| Bone Diseses | Bone (Metabolite) | [SM(d20:1/PGD2)](https://hmdb.ca/metabolites/HMDB0290725) | | NA | HMDB |
| Bone Diseses | Bone (Metabolite) | [SM(d20:1/5-iso PGF2VI)](https://hmdb.ca/metabolites/HMDB0290734) | | NA | HMDB |
| Bone Diseses | Bone (Metabolite) | [SM(d20:1/18:3(9,11,15)-OH(13))](https://hmdb.ca/metabolites/HMDB0290757) | | NA | HMDB |
| Bone Diseses | Bone (Metabolite) | [SM(d18:0/18:0)](https://hmdb.ca/metabolites/HMDB0012087) | | NA | HMDB |
| Bone Diseses | Bone (Metabolite) | [SM(d18:0/18:1(9Z))](https://hmdb.ca/metabolites/HMDB0012089) | | NA | HMDB |
| Bone Diseses | Bone (Metabolite) | [SM(d18:0/23:0)](https://hmdb.ca/metabolites/HMDB0012093) | | NA | HMDB |
| Bone Diseses | Bone (Metabolite) | [17a-Ethynylestradiol](https://hmdb.ca/metabolites/HMDB0001926) |  | NA | HMDB |
| Bone Diseses | Bone (Metabolite) | [Ascorbic acid](https://hmdb.ca/metabolites/HMDB0000044) | | NA | HMDB |
| Bone Diseses | Bone (Metabolite) | [Aluminum](https://hmdb.ca/metabolites/HMDB0001247) | | NA | HMDB |
| Bone Diseses | Bone (Metabolite) | [Vitamin D3](https://hmdb.ca/metabolites/HMDB0000876) |  | NA | HMDB |
| Bone Diseses | Bone (Metabolite) | [Dimethyl sulfoxide](https://hmdb.ca/metabolites/HMDB0002151) | | NA | HMDB |
| Bone Diseses | Bone (Metabolite) | [Prostaglandin D2](https://hmdb.ca/metabolites/HMDB0001403) | | NA | HMDB |
| Bone Diseses | Bone (Metabolite) | [Oxoglutaric acid](https://hmdb.ca/metabolites/HMDB0000208) | | NA | HMDB |
| Bone Diseses | Bone (Metabolite) | [Delta-12-Prostaglandin J2](https://hmdb.ca/metabolites/HMDB0004238) | | NA | HMDB |
| Bone Diseses | Bone (Metabolite) | [Glucosamine](https://hmdb.ca/metabolites/HMDB0001514) | | NA | HMDB |
| Bone Diseses | Bone (Metabolite) | Zinconium | | NA | HMDB |
| Bone Diseses | Bone (Metabolite) | [Thyroxine](https://hmdb.ca/metabolites/HMDB0000248) | | NA | HMDB |
| 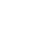<https://hmdb.ca/structures/HMDB0000094/image.svg> Bone Diseses | Bone (Metabolite) | [Polyvidone](https://hmdb.ca/metabolites/HMDB0033843) |  | NA | HMDB |
| Bone Diseses | Bone (Metabolite) | [Citric acid](https://hmdb.ca/metabolites/HMDB0000094) | | NA | HMDB |
| Bone Diseses | Bone (Metabolite) | [Cobalamin](https://hmdb.ca/metabolites/HMDB0002174) | | NA | HMDB |
| Bone Diseses | Bone (Metabolite) | [Thyroxine](https://hmdb.ca/metabolites/HMDB0000248) | | NA | HMDB |
| Bone Diseses | Bone (Metabolite) | [Testosterone](https://hmdb.ca/metabolites/HMDB0000234) | | NA | HMDB |
| Bone Diseses | Bone (Metabolite) | 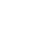<https://hmdb.ca/structures/HMDB0015378/image.svg> [Docetaxel](https://hmdb.ca/metabolites/HMDB0015378) |  | NA | HMDB |
| Bone Diseses | Bone (Diseases) | Retinyl ester | | Blood | HMDB |
| Bone Diseses | Bone (Diseases) | Galactosylhydroxylysine | | Urine | HMDB |
| Bone Diseses | Bone (Diseases) | Galactosylhydroxylysine | | Blood | HMDB |
| Bone Diseses | Bone (Diseases) |  | | Urine | HMDB |
|  |  | Galactosylhydroxylysine | |  |  |
| Bone Diseses | Bone (Diseases) | L-alpha-Aminobutyric acid | | Blood | HMDB |
| Bone Diseses | Bone (Diseases) | Hyperphosphatasia | | Urine | HMDB |
| Bone Diseses | Bone (Diseases) |  | | Urine | HMDB |
|  |  | 25-Hydroxyvitamin D2 |  |  |  |
| Bone Diseses | Bone (Diseases) | Calcium | | Blood | HMDB |
| Bone Diseses | Bone (Diseases) | Phosphate | | Blood | HMDB |
| Bone Diseses | Osteoblast (Diseases) | [12,13-DHOME](https://hmdb.ca/metabolites/HMDB0004705) | | Blood | HMDB |
| Bone Diseses | Osteoblast (Diseases) | [Etidronic acid](https://hmdb.ca/metabolites/HMDB0015210) |  | Blood | HMDB |
| Bone Diseses | Osteoblast (Diseases) | [Prostaglandin A1](https://hmdb.ca/metabolites/HMDB0002656) | | Blood | HMDB |
| Bone Diseses | Osteoblast (Diseases) | [Indoxyl sulfate](https://hmdb.ca/metabolites/HMDB0000682) | | Blood | HMDB |
| Tooth agenesis Diseases | Ectodermal Dysplasia (Metabolite) | [Desmosterol](https://hmdb.ca/metabolites/HMDB0002719) | | Blood, Urine | HMDB |
| Tooth agenesis Diseases | Ectodermal Dysplasia (Metabolite) | [2-Acetamido-2-deoxy-6-O-a-L-fucopyranosyl-D-glucose](https://hmdb.ca/metabolites/HMDB0005817) |  | Blood, Urine | HMDB |
| Tooth agenesis Diseases | Ectodermal Dysplasia (Metabolite) | [N-Acetylgalactosamine 6-sulfate](https://hmdb.ca/metabolites/HMDB0000841) | | Blood, Urine | HMDB |
| Tooth agenesis Diseases | Ectodermal Dysplasia (Metabolite) | [3-Hydroxytetradecanedioic acid](https://hmdb.ca/metabolites/HMDB0000394) | | Blood, Urine | HMDB |
| Tooth agenesis Diseases | Ectodermal Dysplasia (Metabolite) | [8-Isoprostane](https://hmdb.ca/metabolites/HMDB0004659) | | Blood, Urine | HMDB |
| Tooth agenesis Diseases | Ectodermal Dysplasia (Metabolite) | [3-[[5-Methyl-2-(1-methylethyl)cyclohexyl]oxy]-1,2-propanediol](https://hmdb.ca/metabolites/HMDB0036133) |  | Blood, Urine | HMDB |
| Tooth agenesis Diseases | Ectodermal Dysplasia (Metabolite) | [5-Methyltetrahydrofolic acid](https://hmdb.ca/metabolites/HMDB0001396) | | Blood, Urine | HMDB |
| Tooth agenesis Diseases | Ectodermal Dysplasia (Diseases) | alpha-D-Glucose | | Blood, Urine | HMDB |
| Tooth agenesis Diseases | Ectodermal Dysplasia (Diseases) | Bilirubin | | Blood, Urine | HMDB |
| Tooth agenesis Diseases | Ectodermal Dysplasia (Diseases) | [D-Glucose](https://hmdb.ca/metabolites/HMDB0000122) | | Blood, Urine | HMDB |
| Tooth agenesis Diseases | Ectodermal Dysplasia (Diseases) | alpha-D-Glucose | | Blood, Urine | HMDB |
| Tooth agenesis Diseases | Cleidocranial Dysplasia (Metabolite) | [Desmosterol](https://hmdb.ca/metabolites/HMDB0002719) | | Blood, Urine | HMDB |
| Tooth agenesis Diseases | Cleidocranial Dysplasia (Metabolite) | [2-Acetamido-2-deoxy-6-O-a-L-fucopyranosyl-D-glucose](https://hmdb.ca/metabolites/HMDB0005817) |  | Blood, Urine | HMDB |
| Tooth agenesis Diseases | Cleidocranial Dysplasia (Metabolite) | [N-Acetylgalactosamine 6-sulfate](https://hmdb.ca/metabolites/HMDB0000841) | | Blood, Urine | HMDB |
| Tooth agenesis Diseases | Cleidocranial Dysplasia (Metabolite) | [3-Hydroxytetradecanedioic acid](https://hmdb.ca/metabolites/HMDB0000394) | | Blood, Urine | HMDB |
| Tooth agenesis Diseases | Cleidocranial Dysplasia (Metabolite) | [8-Isoprostane](https://hmdb.ca/metabolites/HMDB0004659) | | Blood, Urine | HMDB |
| Tooth agenesis Diseases | Cleidocranial Dysplasia (Metabolite) | [3-[[5-Methyl-2-(1-methylethyl)cyclohexyl]oxy]-1,2-propanediol](https://hmdb.ca/metabolites/HMDB0036133) | | Blood, Urine | HMDB |
| Tooth agenesis Diseases | Cleidocranial Dysplasia (Metabolite) | [Desmosterol](https://hmdb.ca/metabolites/HMDB0002719) | | Blood, Urine | HMDB |
| Tooth agenesis Diseases | Cleidocranial Dysplasia (Metabolite) | [2-Acetamido-2-deoxy-6-O-a-L-fucopyranosyl-D-glucose](https://hmdb.ca/metabolites/HMDB0005817) |  | Blood, Urine | HMDB |
| Tooth agenesis Diseases | Cleidocranial Dysplasia (Metabolite) | [N-Acetylgalactosamine 6-sulfate](https://hmdb.ca/metabolites/HMDB0000841) | | Blood, Urine | HMDB |
| Tooth agenesis Diseases | Cleidocranial Dysplasia (Metabolite) | [3-Hydroxytetradecanedioic acid](https://hmdb.ca/metabolites/HMDB0000394) | | Blood, Urine | HMDB |
| Tooth agenesis Diseases | Cleidocranial Dysplasia (Metabolite) | [8-Isoprostane](https://hmdb.ca/metabolites/HMDB0004659) | | Blood, Urine | HMDB |
| Tooth agenesis Diseases | Cleidocranial Dysplasia (Metabolite) | [3-[[5-Methyl-2-(1-methylethyl)cyclohexyl]oxy]-1,2-propanediol](https://hmdb.ca/metabolites/HMDB0036133) | | Blood, Urine | HMDB |
